# Impact of pre-exposure prophylaxis on HIV-1 drug resistance and phylogenetic cluster growth in British Columbia, Canada

**DOI:** 10.1101/2024.09.06.24313187

**Authors:** Angela McLaughlin, Junine Toy, Vince Montoya, Paul Sereda, Jason Trigg, Mark Hull, Chanson J. Brumme, Rolando Barrios, Julio S.G. Montaner, Jeffrey B. Joy

**Author notes:** **Funding** British Columbia Centre for Excellence in HIV/AIDS, Providence Health Care, Canadian Institutes of Health Research, Public Health Agency of Canada, Genome Canada, and Genome BC.

## Abstract

Oral HIV pre-exposure prophylaxis (PrEP) effectively prevents infection when taken during periods of risk, however, its population-level effectiveness is hindered by incomplete uptake, adherence, and retention. Since PrEP became available free-of-cost in British Columbia (BC), Canada, in January 2018, uptake has been rapid among gay, bisexual, and other men who have sex with men (GBM). Epidemiological evidence suggests adding PrEP onto a background of generalized access to free antiretroviral therapy, under the BC Treatment as Prevention (TasP) strategy, had a synergistic effect on reducing new infections. Here, we sought to evaluate the impact of PrEP on HIV transmission and drug resistance in phylogenetic clusters. In a retrospective cohort study, we evaluated whether baseline HIV drug resistance and phylogenetic clustering were more likely among newly diagnosed PrEP users in BC (n=39) compared to non-PrEP users (n=566) during the same diagnosis period from October 23, 2018 to December 5, 2022. Subsequently, we evaluated heterogeneity in PrEP-related reductions of the effective reproduction number (R_e_) across BC, key populations, and phylogenetic clusters. Stochastic branching processes of phylogenetic clusters informed by R_e_ preceding PrEP were used to estimate diagnoses averted via PrEP. Newly HIV diagnosed PrEP users were significantly more likely than non-PrEP users to join phylogenetic clusters and carry baseline nucleoside-analogue reverse transcriptase inhibitor (NRTI) resistance-associated mutation M184I/V. Despite reductions in growth rate and R_e_ in the GBM population overall and in 50% of active GBM-predominant clusters following PrEP availability, we highlight predominantly GBM and PWID clusters with high or increasing R_e_. Across active phylogenetic clusters and non-clustered new diagnoses, we estimate PrEP averted approximately 20 new HIV diagnoses per year in BC since 2018. These findings highlight how PrEP has reduced HIV burden, while illuminating groups that could benefit from prioritized PrEP education, access, and retention.

## Introduction

Optimal control of HIV/AIDS requires equitable delivery of antiretroviral treatment (ART) to all people living with HIV (PLWH) immediately following HIV diagnosis and prevention interventions, including pre-exposure prophylaxis (PrEP) to populations at elevated risk, informed by longitudinal epidemiological monitoring, including via phylogenetic identification of HIV transmission clusters. Although HIV morbidity, mortality and incidence have declined in well-managed epidemics in the developed world, including in British Columbia (BC), Canada,^1^ ongoing transmission continues to disproportionately affect some key populations.

Oral PrEP effectively prevents HIV-1 acquisition when taken during periods of risk among gay, bisexual, and other men who have sex with men (GBM),^2^ transgender women (TGW),^2^ heterosexuals,^3^ and people who inject drugs (PWID).^4^ Barriers to access, racial disparities,^5^ limited eligibility or coverage, inconsistent adherence,^6^ and program non-retention^7^ have resulted in sub-optimal oral PrEP effectiveness. In 2016, Health Canada approved once-daily oral PrEP, Truvada^®^, comprised of tenofovir disoproxil fumarate (TDF)/emtricitabine (FTC) for adults at elevated risk of HIV acquisition, including GBM and serodiscordant heterosexual couples,^8^ followed by generic TDF/FTC in 2017.^9^

Since January 1, 2018, PrEP has been fully publicly funded and centrally distributed for BC residents with greater likelihood of HIV acquisition as determined by local PrEP guidelines.^10^ PrEP uptake in BC has expanded rapidly, with 9737 cumulative participants by June 30, 2022,^7,11^ consisting of 97·0% cis GBM, 1·3% TGW, 0·9% cis-women, and 0·5% transgender men (TGM).^7^ Non-cisgender men (TGM, TGW, cis-women) were less likely to persist with PrEP, as were those who were younger and had no prior PrEP use.^7^ Differences in PrEP awareness, uptake, access, and persistence manifest as differential reductions in HIV transmission. Lapsed prescriptions (greater than 6 months beyond expected refill date based on daily PrEP use) and incomplete adherence have resulted in 39 newly diagnosed HIV infections in PrEP users between October 23, 2018 and December 5, 2022 (0·4% of PrEP participants).

Population-level effectiveness of PrEP has been estimated through cohort studies, simulations, and mathematical models, but not often at the phylogenetic cluster level. In a compartmental model of HIV in BC, prioritized provision of PrEP to GBM with higher likelihood of HIV acquisition, in combination with Treatment as Prevention (TasP), was associated with reduced incidence and R_e_ below one in susceptible GBM.^12,13^ Clustering the BC population by reported potential HIV exposure, testing behaviour, and PrEP interest increased the estimated population-level effectiveness of PrEP.^14^ In the Australian EPIC-NSW cohort study, PrEP availability resulted in a 25% relative reduction of new HIV diagnoses in GBM.^15^ In South Africa, prioritizing PrEP to groups with higher likelihood of HIV acquisition was more efficient than based on partnership alone.^16^ A stochastic mechanistic model suggested PrEP averted 20% of HIV acquisitions from 2015-2021 in Montreal.^17^ Expanding oral PrEP coverage to 80% was estimated to avert 8% of new HIV infections in Australia, 15% in Thailand, and 26% in China over 40 years.^18^ In the US, network-based mathematical models were applied to estimate that 33% of cases over the next decade could be averted with 40% coverage of MSM and 62% adherence.^19^ No HIV seroconversions were reported over 850 person-years of PrEP use in Northern California, however there were two seroconversions in individuals who had discontinued PrEP use,^20^ highlighting that PrEP effectiveness relies on long-term use and support.

Pre-treatment drug resistance, either through the transmission of resistant variants, or acquired resistance due to the use of TDF/FTC during undiagnosed or acute HIV infection, adversely impacts clinical outcomes by limiting treatment options.^21–23^ Mutations conferring resistance to NRTI drugs in PrEP, TDF (K65R) and FTC (M184V/I), were identified in individuals retrospectively found to have acute HIV infections who were prescribed PrEP.^24^ In New York, past and recent PrEP use were associated with significantly elevated risk of baseline M184I/V, adjusted for transmission risk group.^21^ In a meta-analysis, individuals exposed to PrEP during acute infection were significantly more likely to have TDF or FTC resistance.^25^ HIV acquisition during PrEP use has occurred in randomized control trials, but is confounded by incomplete adherence and reporting bias.^24^ Evaluating HIV drug resistance in phylogenetic trees can illuminate transmission of drug resistance in expanding phylogenetic clusters, routinely monitored for HIV surveillance in BC,^26^ which can represent groups linked by recent transmission events.

In this study, we sought to estimate impacts of PrEP use on baseline drug resistance and phylogenetic clustering among new HIV diagnoses, and the population-level impacts of PrEP on the effective reproductive number (R_e_) of phylogenetic clusters. We tested the hypotheses that i) newly diagnosed PrEP users were more likely to cluster than non-PrEP users, under the expectation that PrEP eligibility and use is related to behaviour associated with potential exposure and linkage to sexual transmission networks, and that ii) they have elevated baseline NRTI resistance, under the expectation that incomplete PrEP adherence led to suboptimal drug concentration during acute infection. We further hypothesized that iii) cluster-specific R_e_, and diagnoses averted have been heterogeneous since PrEP availability, amid broad R_e_ reductions in the BC GBM population.

## Methods

### Study design

Data for this retrospective cohort study were obtained on February 9, 2023, representing data up to December 31, 2022, from the Drug Treatment Program (DTP) at the BC Centre for Excellence in HIV/AIDS (BC-CfE) representing PLWH connected to publicly-funded treatment in BC, Canada. Participant data were de-identified and sequences were assigned random six-character identifiers. Sequencing was performed by clinical staff at the BC-CfE and genotypic data were stored in the BC-CfE access-controlled facility within a secure, encrypted Oracle database. Investigators had no access to nominal data. Research ethics were approved by University of British Columbia-Providence Health Care Research Institute (REB #H20-02859) on December 7, 2020 and amended September 8, 2022.

### Participants

We evaluated a subset of 10,740 participants with HIV sequences in the DTP including 39 newly diagnosed PrEP users, with dates of first detectable plasma HIV viral load (VL), used as a proxy for diagnosis, ranging from October 23, 2018 to December 5, 2022, and a control group of 566 non-PrEP users newly diagnosed over the same period. Individuals enrolled for PrEP who never filled prescriptions were not considered PrEP users. New participants with previous ART experience (PLWH who have immigrated) were excluded.

### Procedures and outcomes

De-identified participant data included viral genetic sequences, gender, sex-at-birth, age, residence census tract and regional health authority, prescriber census tract, date of first VL, date of first ART, ART prescriptions, PrEP dispensations (if applicable), self-reported potential HIV exposures at DTP enrolment i.e., key populations (GBM; PWID; heterosexual (HET); blood; Hepatitis C virus; other), VL, baseline CD4+ cell count, history of acquired immune deficiency syndrome (AIDS) diagnosis, and if applicable, date and cause of death.

We analyzed 41,941 HIV-1 partial pol (*protease* and partial *reverse transcriptase*) sequences from 10,740 DTP participants collected between May 30, 1996 and December 31, 2022 (cleaning, alignment, and subtype assignment in **Supplementary**) and 8739 *integrase* (int) sequences from 4052 patients (1-35 sequences per patient). Sequences from newly diagnosed individuals were screened for resistance-associated mutations (RAMs) using the Stanford HIV Drug Resistance Database (HIVdb) version 9.6.^27^

A set of shuffled alignments were generated to infer 100 bootstrap approximate maximum likelihood (ML) phylogenetic trees in FastTree 2.1 with a generalized time reversible (GTR) substitution model.^28^ Trees were outgroup rooted on the oldest subtype B sequence. Phylogenetic clusters comprised a minimum of five members with viruses sharing pairwise patristic distance <0·02 substitutions per site in greater than 90% of bootstraps.^26^ We evaluated the effect of rooting using outgroups, midpoint, and root-to-tip regression on the distribution of cluster sizes (**Fig. S3**). Separate ML trees were inferred for key clusters with IQ-TREE under a GTR substitution model and 1000 resamples for ultrafast bootstrap support values,^29,30^ and rooted using root-to-tip regression optimized by residual mean squares in ape (**Fig. S8-15**).^31^ We calculated annual HIV incidence and prevalence per 100,000 (**Fig. S16**) for 14,919 DTP participants (with or without a sequence available) up to December 31, 2022, adjusted by quarterly population size.^32^

The prevalent PLWH population size was calculated as cumulative new cases minus deaths and emigrants by quarter; and growth rate as new cases per prevalent case per quarter.

Diagnoses over time were used to estimate incidence,^33^ and the instantaneous effective reproduction number (R_e_) in BC, key populations, and active phylogenetic clusters (with at least one new diagnosis since 2018) with at least 10 members (inadequate data for clusters sized 5-10) using EpiEstim with a gamma-distributed serial interval.^34^ Parameters compared for R_e_ estimation in BC (**Figs. S19-22**), key populations (**Figs. S23-24**), and clusters (**Figs. S25-28**) included serial interval mean of 0.5 year (y), 1 y, 2 y, or 5 y, and standard deviation (sd) of 0·5 y, 1 y, or 2 y; estimation interval of 0·25 y, 0·5 y, or 1 y; and R_e_ smoothing over 30 days (d), 90 d, or 365 d. Primary analyses represent mean 1 y, sd 0.5 y, estimation interval 0.5 y, and 90 d smoothing. We fitted average R_e_ in the periods before PrEP (Jan. 2016 – Dec. 2017), during PrEP and before COVID-19 (Jan. 2018 - Feb. 2020), during PrEP and COVID-19 (Mar. 2020 – Feb. 2022), and during PrEP and post-COVID-19 (Mar. 2022 - Dec. 2022). Fold-change (FC) in R_e_ with PrEP (‘PrEP effect’) was calculated as the ratio of R_e_ during PrEP and before COVID-19 over R_e_ before PrEP in key populations and clusters.

Projected lifetime ART cost savings were estimated by multiplying cases averted by the average lifetime cost of treating a PLWH in Canada ($250,000).^35^ This does not constitute a cost-effectiveness analysis, as we do not compare the costs of PrEP.^41^

### Statistical analyses

Characteristics associated with PrEP use among newly diagnosed were evaluated with chi-square tests or medians were compared using Kruskal-Wallis (**Table S4**), as were factors associated with clustering stratified by PrEP use (**Table S5**). We tested the hypothesis that newly diagnosed PrEP users had higher clustering proportion and higher proportion of baseline NRTI resistance than non-PrEP users with chi-squared tests if greater than 80% of expected frequencies exceeded five, otherwise Fisher’s exact tests were applied. Lineage-level viral diversification rates in newly diagnosed with or without PrEP were compared using a Kruskal-Wallis test (**Supplementary**). Proportions of newly diagnosed with and without PrEP use who had baseline drug resistance mutations were compared using Fisher’s exact tests with p-values adjusted using the Benjamini-Hochberg method.^36^

Stochastic branching processes were used to simulate HIV cluster growth with and without PrEP, whereby the number of secondary cases followed a negative binomial distribution defined by mean R_e_ and dispersion k.^37–39^ More details in **Supplementary**. We assumed a gamma-distributed serial interval (mean 1 y, sd 0.5 y); k between 0.1 and 0.3; and R_e_ as the observed cluster R_e_ (to recapitulate observed cases with PrEP) or counterfactual cluster R_e_ adjusted by PrEP effect to simulate absence of PrEP. Seeds were adjusted to align the number of new samples observed and simulated using observed R_e_ (**Table S3**). For each cluster, 4000 simulations were run from January 1, 2018 to January 1, 2023. Averted diagnoses and samples were calculated as the difference in mean and bootstrapped 95% CI of simulations with and without PrEP. Poisson generalized linear models were evaluated to identify cluster characteristics associated with diagnoses averted. Factors evaluated for inclusion in the model included cluster size in 2017 (immediately preceding PrEP availability) and among cluster members, percentage in key populations, percentage residing in each BC health authority, median age in 2023 when the study was conducted, median age at first ART, and percentage of new diagnoses with PrEP use.

## Results

### Clustering of newly diagnosed PrEP users

Newly HIV diagnosed PrEP users (n=39) in BC were significantly more likely than newly diagnosed non-PrEP users (n=566) to be male (100% vs. 79·5%; chi-squared test, p=0·011; **Table S4**), GBM (97·1% vs 56·7% of reported, p<0·001), reflecting that the majority of PrEP users are GBM, younger at first ART (median 32 vs. 37 years old; Kruskal-Wallis test, p=0·033), and have a higher baseline CD4+ T-cell count (490 vs. 380, p=0·003).

By December 31^st^, 2022, there were 246 BC HIV phylogenetic clusters (comprising at least 5 individuals with viruses sharing pairwise patristic distance less than 0·02 substitutions per site in at least 90% of bootstraps), 84 of which were active, with at least one new diagnosis since 2018. Tree method negligibly affected cluster identification (**Fig. S3**). All clusters joined by PrEP users were also joined by at least one non-PrEP user (**Fig. S1**). Newly diagnosed PrEP users were significantly more likely to cluster than non-PrEP users (chi-squared test p=0·0075, **Fig. S2**). Of 39 newly diagnosed PrEP users, 30 joined 13 unique clusters (76·9% clustered, one-group proportions test 95% confidence interval (CI): 60·3-88·3%), compared to 303 of 566 non-PrEP users (53·5% clustered, 95% CI: 49·3-57·7%), who joined 84 unique clusters (**Table S6**). Among GBM, 82.4% of newly diagnosed PrEP users clustered, compared to 54.7% of non-PrEP users (**Table S5**). Newly diagnosed PrEP users also had significantly higher lineage-level viral diversification rates, indicative of rapid phylogenetic branching events, than non-PrEP users (**Fig. S2**). In a binomial model, newly diagnosed with PrEP use had 2·6-times (95% CI: 1·2-6·4) higher odds of clustering compared to non-PrEP users, adjusted for GBM, age at first ART and baseline CD4 count.

### Comparison of baseline drug resistance among newly diagnosed by PrEP use

Drug resistance mutations were identified and resistance scores were calculated using Stanford HIVdb version 9·6.^40^ Differences in prevalence of baseline resistance-associated mutations (RAMs) between newly diagnosed PrEP users and non-PrEP users were evaluated by drug class (**Fig. S5**) and mutation (**Fig. 1**, **Table S7**), acknowledging that some RAMs like T215S and K219N do not on their own confer clinically-relevant drug resistance levels. There were no significant differences in the proportion of newly diagnosed PrEP users and non-PrEP users with baseline RAMs to any drug class (chi-squared test, p=0·12), nor RAMs of specific drug classes (NNRTI, Fisher’s exact test: p=0·12; NRTI, p=0·26; PI, p=0·2 (**Fig. S5**). Among 39 newly diagnosed PrEP users, there were 14 individuals with any RAMs, six with NNRTI RAMs, eight with NRTI RAMs, and two with PI RAMs. There were no baseline integrase strand inhibitor (INSTI) RAMs among newly diagnosed PrEP users, although five non-PrEP users had baseline INSTI RAMs (**Fig. 1**).

**Fig. 1.**
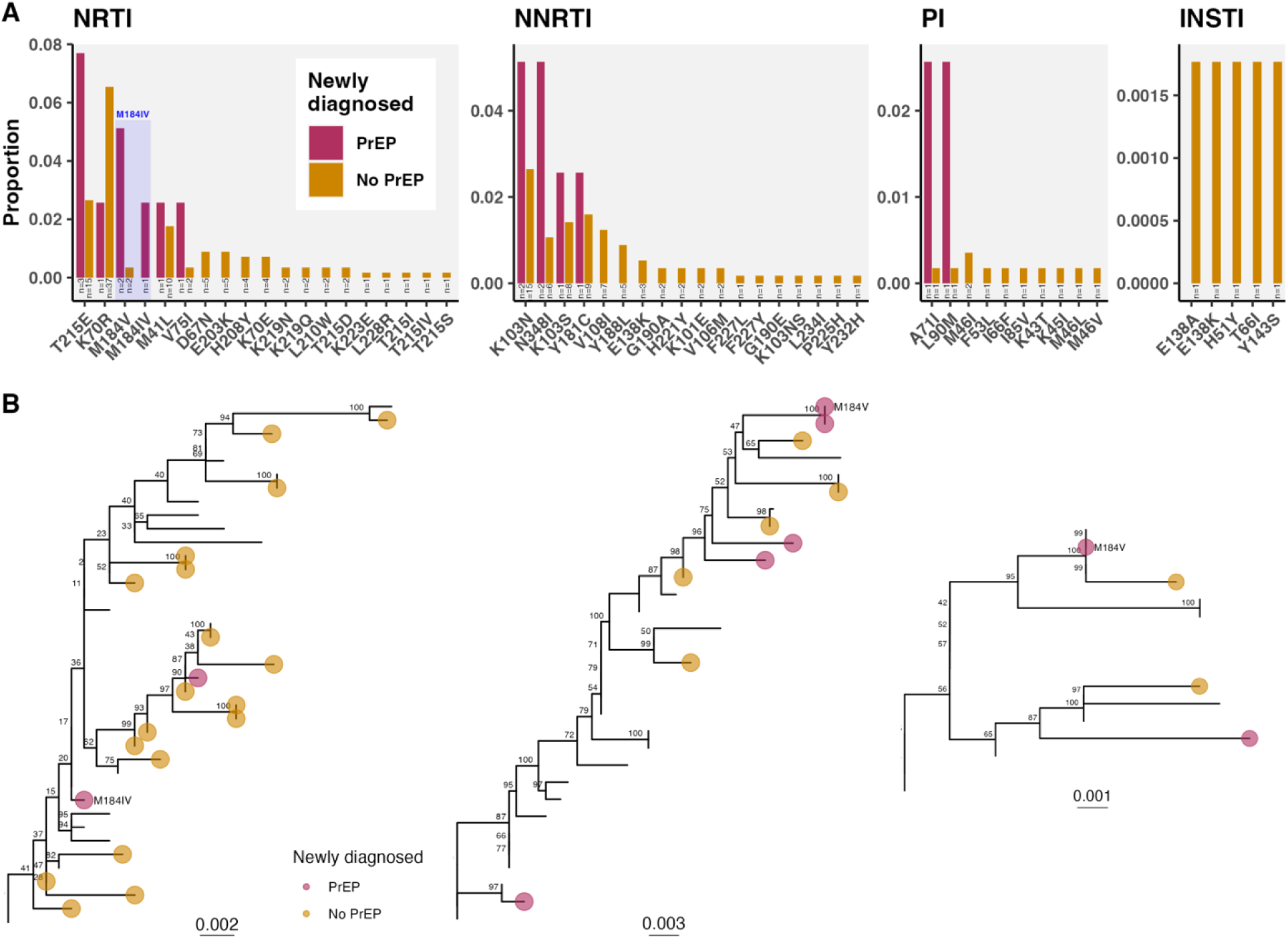
Baseline HIV drug resistance in newly diagnosed individuals with and without PrEP. **A)** Proportion of newly diagnosed with or without PrEP use with baseline drug resistance mutations to NRTI, NNRTI, PI, and INSTI drug classes. Annotations indicate number of individuals with baseline resistance. Mutation M184I/V shaded in blue. **B)** Clustered viruses with baseline M184V or M184I/V. Subtrees pruned from maximum likelihood trees inferred for individual clusters in IQTREE.

### PrEP use associated with elevated probability of baseline NRTI drug resistance

Baseline NRTI resistance to FTC, lamivudine (3TC), and abacavir (ABC), conferred by M184V or M184I/V, was more frequent in newly diagnosed PrEP users than non-PrEP users (**Fig. 1**). Three of 39 newly diagnosed PrEP users (7·7%) had baseline M184V or M184I/V, compared to 2 of 566 (0·35%) non-PrEP users (Fisher’s exact test, adjusted p-value=0·025; **Table S7**; power=79% at alpha=0·05). M184I/V was the only baseline RAM significantly more common in PrEP users than non-PrEP users (**Table S7**).

Newly diagnosed PrEP users with baseline M184V had the two lowest proportion of days covered by PrEP (0·60 and 0·63 vs. median=0·88 across newly diagnosed PrEP users) and had significantly shorter time from last PrEP dispensation to first detectable VL (76 d vs. 373 d; Kruskal-Wallis test, p=0.045). The newly diagnosed PrEP user with M184I/V had a PrEP prescription filled 29 days before first detectable VL. It was later identified that this was a clinical miss, as the individual had acute HIV seroconversion at PrEP initiation and were diagnosed at one month follow-up. Nearest phylogenetic neighbours to viruses with M184I/V did not have M184I/V (**Fig. 1B**).

### Transmission of resistance-associated mutations in phylogenetic clusters

There were 28 active clusters with at least one new diagnosis since 2018 with baseline resistance mutations (**Fig. S6**). NRTI RAM, K70R, and NNRTI RAM, K103N, were increasingly detected at baseline (**Fig. S4**) and transmitted within clusters (**Fig. S6, Fig. S13**). NRTI RAMs detected multiple times within clusters, suggesting possible transmission include thymidine analogue mutation (TAM) revertant T215E (100% of 14 new cases in cluster 114, **Fig. S13**), K70R (27.3% of 11 new cases in cluster 49, **Fig. S10**; 100% of 2 new cases in cluster 185; and outside clusters, **Fig. S6**), D67N (11.5% of 26 new cases in cluster 137, **Fig. S14**), and M41L (4.1% of 94 new cases in cluster 31, **Fig. S9**; present in clade of cluster 137, **Fig. S14**). Baseline NNRTI RAMs detected multiple times within clusters included K103N (2.1% of 94 new cases in cluster 31, **Fig. S9**; 80% of 5 new cases in cluster 97; 40% of 5 in cluster 194), K103S (83.3% of 6 new cases in cluster 234), Y181C (100% of 5 new cases in cluster 197), and N348I (85.7% of 7 new cases in cluster 41). No baseline K65R, conferring resistance to tenofovir, was detected among new cases within or outside clusters.

### Provincial HIV dynamics following PrEP

Within the DTP, representing PLWH connected to publicly-funded treatment in BC, HIV incidence declined from 9.2 new cases per 100,000 in 2012 to 2·7 new cases per 100,000 in 2022 (**Fig. S16**). Before PrEP, new HIV cases overall decreased annually by -12·1% in 2016 and -23·4% in 2017 (**Fig. 2**), with most rapid declines among PWID (-19·7% in 2016, -49·0% in 2017), but less so among GBM (-4·7% in 2016, -11·4% in 2017, **Fig. S18**). Following PrEP, annual new HIV cases in GBM declined -19·2% in 2018 and -8·9% in 2019, and GBM accounted for fewer new cases. During COVID-19, new HIV cases in GBM declined -26·1% in 2020, whereas they increased 6·4% among HET. Post-COVID, new HIV cases increased by 5·7% in GBM in 2022 likely due to increased potential exposures and testing, despite lowering -54·1% in PWID and -28·3% in HET. Among GBM, average R_e_ was around 1 before PrEP, then dropped below 1 during PrEP, and further below 1 during COVID-19 (**Fig. 2, Fig. S23**). R_e_ among PWID (**Fig. S24**) and HET were elevated following PrEP, increased during COVID-19, and have been relatively low post-COVID-19.

**Fig. 2.**
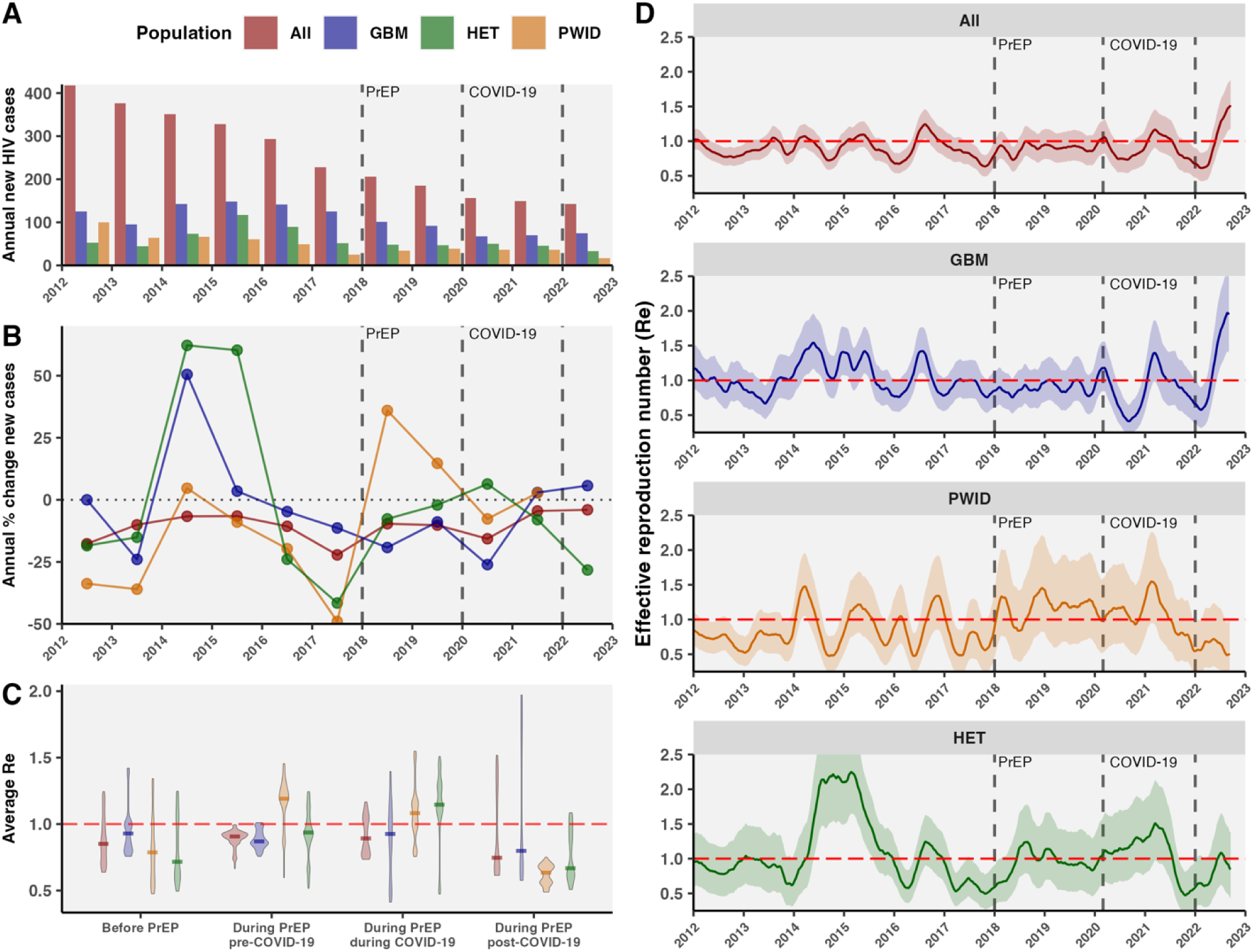
HIV dynamics in BC from 2015 to 2023 in the context of PrEP and COVID-19. A) Annual new HIV diagnosed cases (based on first ART date) overall and among key populations, GBM, HET, and PWID. **B**) Annual percentage change in new HIV diagnoses. **C)** Average effective reproduction number (R_e_) for periods before PrEP (2016-2017), during PrEP and before COVID-19 (2018-Feb. 2020), during PrEP and during COVID-19 (Mar. 2020-2021), and during PrEP and post-COVID-19 (2022). **D)** Temporal dynamics of R_e_ overall and in key populations.

### Differential cluster growth following availability of PrEP

We evaluated heterogeneity in R_e_ dynamics in phylogenetic clusters following PrEP. There were 84 active clusters with at least one new case since 2018; 53 clusters had more than 10 members by Feb. 2023 (**Fig. S1, Fig. S3**). Of active clusters, 52 were predominantly GBM, 30 PWID, and 2 HET (**Table S8**).

Transmission within and outside BC HIV phylogenetic clusters was variably impacted by PrEP (**Fig. 3, Fig. S25-30**). Cluster 31, the largest GBM-predominant cluster (size=391, 89% GBM, 94 new cases since 2018), had stable R_e_ near 1, followed by an increase in mid-2017, then a decline since PrEP availability (fold-change (FC) R_e_ with PrEP<1). Other GBM-predominant clusters (cluster 95, size=158, 27 new cases; and cluster 22, size=42, 19 new cases) had sustained declines through PrEP, with recent resurgences post-COVID-19 (**Fig. S30**). The two largest PWID-predominant clusters (clusters 49 and 57, sizes 459 and 471, 91% and 86% PWID) have had limited new cases since 2018 (11 and 19). R_e_ of PWID cluster 57 was stably below or near 1 from 2015 to 2018, then rose significantly above 1 in 2019, coming down in 2020 before rising above 1 post-COVID-19. PWID cluster 137 (size=90, 26 new cases, 88% PWID) has also had volatile R_e_, peaking in 2017-2018, dropping in 2019, and rising again in late 2019 and 2020. Comparing the PrEP effect on cluster-level R_e_ in key populations, 50% of GBM clusters had reduced R_e_ following PrEP, compared to 33·3% of PWID clusters (**Fig. 3**).

**Fig. 3.**
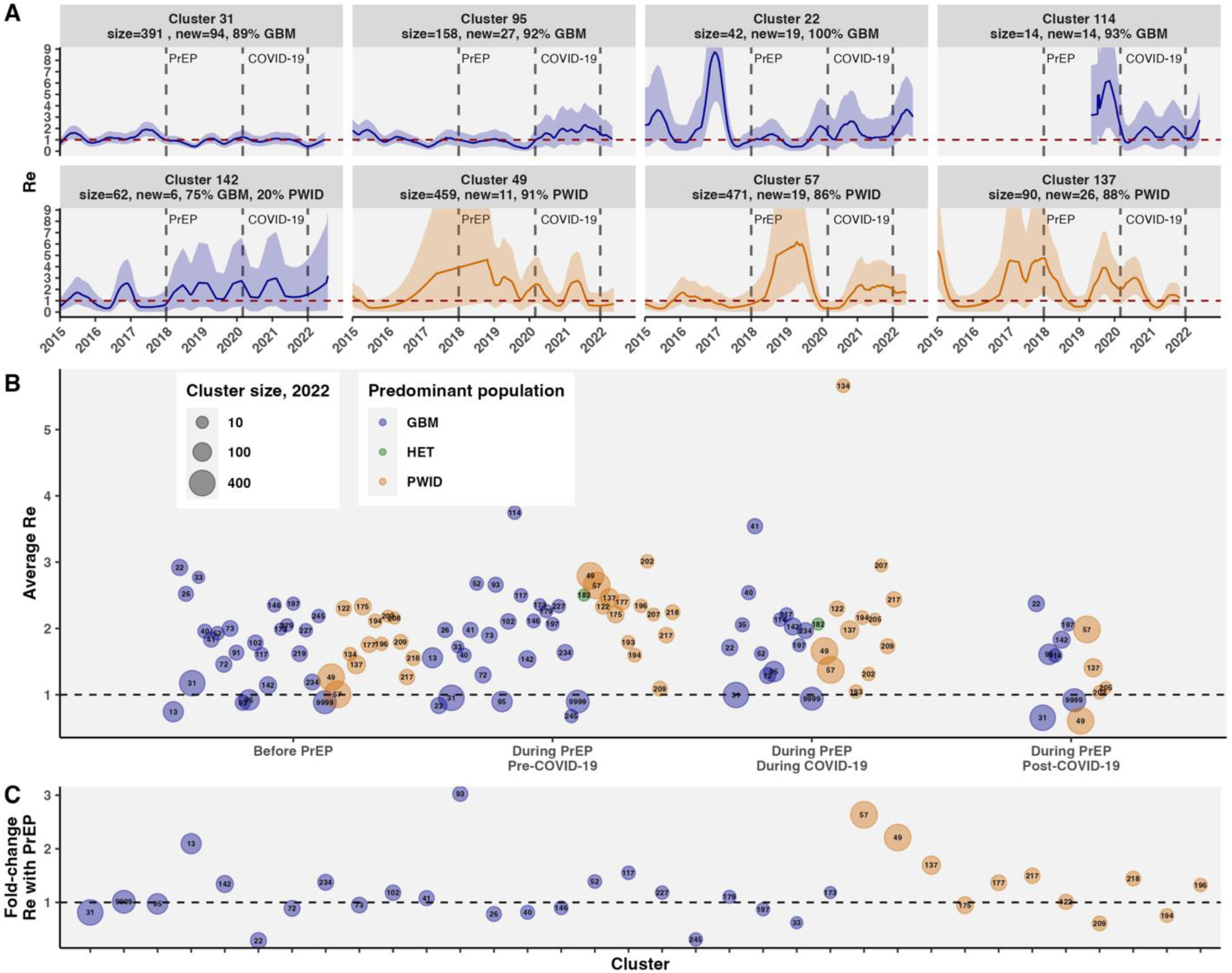
HIV phylogenetic cluster reproduction number in the context of PrEP and COVID-19. A) Phylogenetic cluster-level effective reproduction number (R_e_) for eight key active clusters in BC from 2015 to 2022. R_e_ estimated using EpiEstim with gamma-distributed serial interval with mean 1 y and sd 0.5 y, estimating window 0·5 y, and 90 d smoothing. **B)** Piecewise average R_e_ for active clusters in periods before PrEP, during PrEP and before COVID-19, during PrEP and during COVID-19, and during PrEP and post-COVID-19. Annotated by arbitrary cluster identifier, colored by predominant risk group, and area by cluster size in 2022. **C)** Fold-change (during/before) in active clusters’ R_e_ following PrEP. Clusters sorted on the x-axis by predominant risk group and size in 2022.

### Diagnoses averted via PrEP in phylogenetic clusters estimated with stochastic branching processes

Diagnoses averted in phylogenetic clusters via PrEP between 2018 and 2022 were estimated using stochastic branching processes (**Table S3, Fig. S32-34**).^37–39^ Diagnoses averted were calculated as the difference between new diagnoses without and with PrEP simulated with cluster R_e_ specified as observed R_e_ (with PrEP) or adjusted R_e_ (counterfactual without PrEP).

PrEP averted the most diagnoses in large and medium sized GBM-predominant clusters, including clusters 31, 22, 245, and 33 (**Fig. 4, Fig. S34**). In cluster 31, we estimated 86·8 new diagnoses (95% CI 84·2-89·4) from 2018-2022 in simulations based on observed R_e_ (with PrEP, **Fig. S32**), consistent with the observed 94 new sequenced diagnoses; without PrEP, we estimated up to 115·7 (112·3-119·1) new diagnoses (**Fig. S34**), amounting to 28·9 (28·1-29·7) diagnoses averted in cluster 31 in five years. In cluster 22, there were 24·0 (21·7-26·4) new diagnoses with PrEP (consistent with 19 new diagnoses since 2018), compared to 49·9 (45·9-54·1) without PrEP, amounting to 25·9 (24·2-27·7) diagnoses averted. PrEP was also associated with reduced R_e_ in two medium-sized, relatively young, GBM-predominant clusters with one new diagnosis since 2018 (cluster 245, size 2023=13, median age=34; cluster and 33, size 2023=10, median age=37), dropping from R_e_ well above 1 before PrEP to below 1 during PrEP. We estimated 20·0 (17·9-22·1) diagnoses averted in cluster 245 and 6·0 (5·2-64·0) diagnoses averted in cluster 33. In total, among active phylogenetic clusters in BC with HIV diagnoses averted, we estimated 107·6 (99·1-115·7) diagnoses averted via PrEP in BC between Jan. 2018 and Jan. 2023, or 21·5 (19.8-23.1) diagnoses averted per year. Clusters with negative diagnoses averted represent missed opportunities for PrEP and include PWID-predominant clusters 137, 205, and 49, as well as non-clustered cases (cluster 9999, predominantly GBM but mixed) and GBM-predominant clusters 142, 234, and 13. Cluster characteristics univariately associated with fewer diagnoses averted include higher median age, lower proportion of new diagnoses with PrEP access, and living outside of Vancouver Coastal Health Authority. For instance, cluster 142 is comprised of relatively older individuals (median age=55, median age first ART=43), with a majority of new diagnoses in Vancouver Island Health Authority (**Table S8**). In a Poisson model of diagnoses averted, clusters with higher median age and lower % GBM had significantly fewer diagnoses averted, adjusted for cluster size at the end of 2017 and % residing in Vancouver Coastal Health Authority (**Table S9**).

**Fig. 4.**
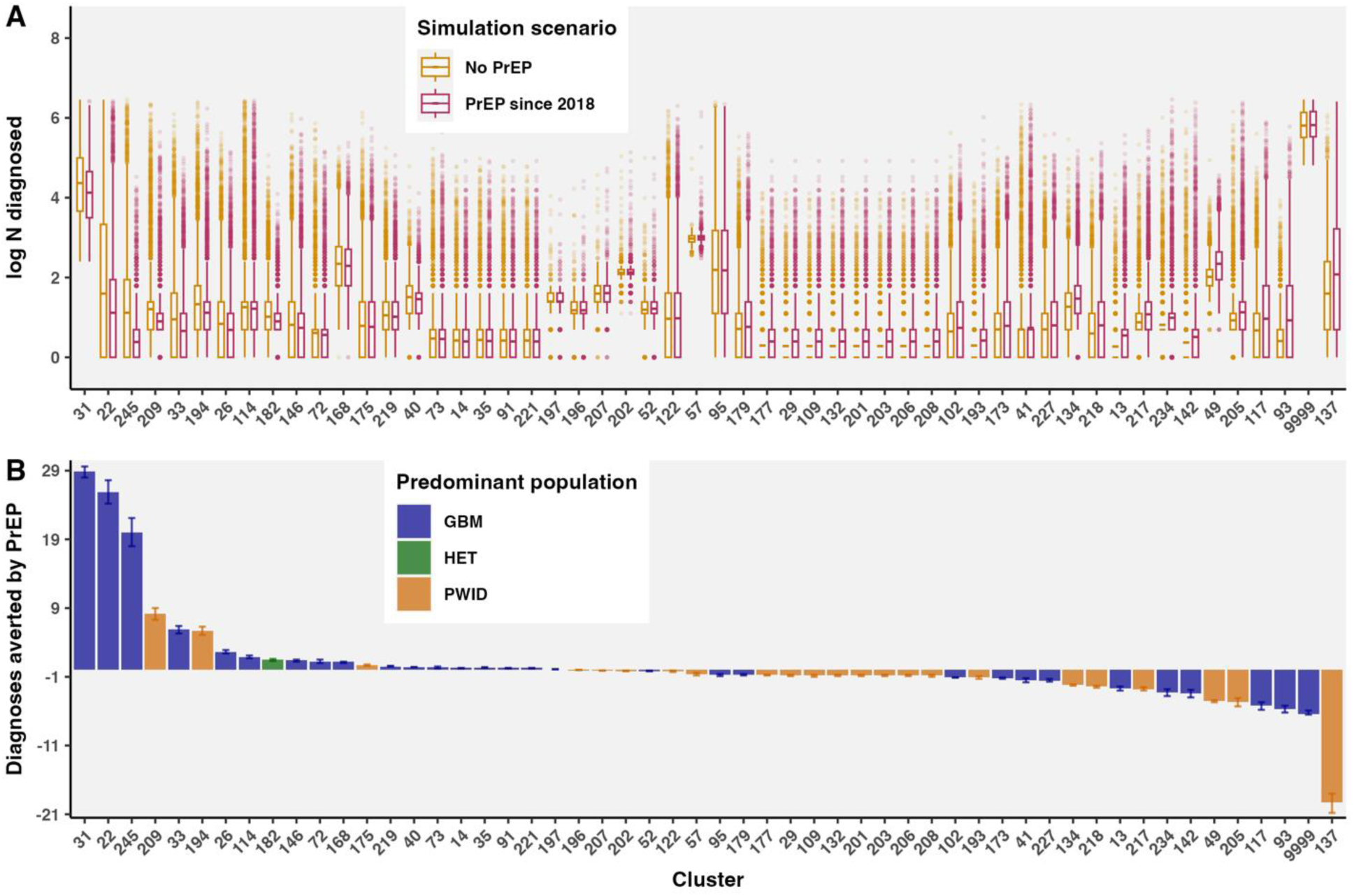
Diagnoses averted via PrEP in active phylogenetic clusters estimated using stochastic branching processes. A) Simulations of new diagnoses in clusters from January 2018 to January 2023 were run with observed cluster R_e_ (with PrEP since 2018) or with R_e_ adjusted by the estimated PrEP effect (no PrEP counterfactual scenario). **B)** Diagnoses averted by PrEP from 2018 to 2022 by cluster, estimated as the difference between new diagnoses in simulations with and without PrEP. Bar height represents the mean and error bars represent the bootstrap 95% CI across simulations.

Assuming the overall lifetime cost associated with each HIV diagnosis is $250,000 Canadian dollars,^35^ 107·6 (99·1-115·7) diagnoses averted via PrEP in BC between Jan. 2018 and Jan. 2023 represents an averted lifetime treatment cost of $26,900,000 Canadian dollars ($24,775,000 - $28,925,000). However, cost-effectiveness analyses must weigh potential benefits of prioritized PrEP programs against the costs including drugs (estimated as $360 USD per person per year in the US), clinic visits ($104 USD), diagnostic tests, and administrative hours.^41^

## Discussion

Our analyses suggest publicly funded PrEP in BC, layered on top of generalized access to free ART under the TasP strategy, has averted more than 100 new diagnoses since 2018, concentrated in active GBM clusters, resulting in substantial reductions in morbidity and mortality. While PrEP has dampened transmission in large GBM-predominant clusters, other clusters had fewer cases averted, illuminating treatment and prevention gaps for future public health interventions.

Newly diagnosed PrEP users were more likely to cluster than non-PrEP users, which could reflect systematic differences between health-seeking and risk-taking behaviour among those who did and did not access PrEP before seroconverting. We highlighted fewer cases averted via PrEP in clusters with higher median age, lower proportion GBM, and low proportion of previous PrEP use. PrEP uptake, adherence, and retention differ by age, risk perception,^42^ geography, social norms, substance use, ease of refills, community support, and stigma. Newly diagnosed PrEP users represent missed opportunities for program retention through better tools for support, such as peer workers or long-acting options. Whereas newly diagnosed non-PrEP users represent a missed opportunity for PrEP education and access. We further highlight that contacts of members of active phylogenetic clusters represent ongoing opportunities for PrEP education, eligibility, and access. Among HIV-negative GBM not using PrEP in Vancouver in 2018, 28% were uncomfortable asking doctors for PrEP, highlighting the importance of educating physicians as PrEP providers,^42^ and exploring potential for alternative care delivery models such as online PrEP services to improve PrEP access and retention.

Newly diagnosed PrEP users were more likely to have baseline M184I/V conferring reduced susceptibility to NRTIs, FTC, 3TC, and ABC, and individuals with M184I/V had a short time since last PrEP dispensation. This suggests inadvertent PrEP exposure during acute infection may have strongly selected for M184V, or prevented the reversion of M184I/V-containing variants, potentially limiting treatment options with regimens containing 3TC or FTC.^43^ Baseline M184I/V was found among individuals with more recent PrEP dispensations and in one instance occurred in an individual with acute HIV infection (false negative) during PrEP initiation. Phylogenetic neighbours of PrEP users with baseline M184V did not carry the mutation, suggesting it was acquired *de novo*, although we cannot exclude the possibility it may have been an undetected minority variant or had reverted to wild-type prior to testing in these individuals. PrEP clients likely undergo more frequent HIV diagnostic testing, raising the possibility of earlier enrolment in the DTP and earlier drug resistance testing, which could impact the likelihood of detecting M184V. We lack evidence of onward transmission of M184I/V, corroborating its fitness cost by the time baseline drug resistance testing occurs.

In the Swiss HIV epidemic, M184V along with RAMs, D67N, K70R, and K219Q conferred negative fitness, while L90M was advantageous.^44^ In the absence of drug selection pressure, M184V was rapidly lost in a UK cohort, whereas T215 revertants, D67N, L210W, and K219Q/N persisted.^45^ We reported probable transmission of RAMs within clusters with clinical implications, as K70R reduces susceptibility to AZT and K103N reduces susceptibility to nevirapine and efavirenz.^27^ Detection of T215E TAM revertant mutation, in 100% of 14 new cases within a key cluster could suggest transmission of a resistant variant that is in the process of reverting to wild-type.

R_e_ estimates depend on the serial interval, i.e., the time between successive infections’ symptom onset.^46^ Compared to viruses with short period of infectiousness, HIV serial intervals can vary widely, as transmission could occur during acute infection (45% of transmissions in the first year in a Detroit MSM cohort),^47^ or later if there is viral rebound. We explored realistic serial intervals, settling on 1 y for the primary analysis under the premise of widely available testing and treatment in BC. Our estimates assume that serial interval is comparable across clusters and key populations. The cluster-specific R_e_ confidence intervals and uncertainty were wide when newly diagnosed cluster members were sparse (Fig. 3, Fig. S25).

Although we filtered estimates with CI width greater than 10, a more stringent cut-off or wider estimating window may have been warranted in some instances. Reducing noise in time-series data could improve R_e_ estimates for low incidence,^48^ which we circumvented with wide estimation windows and smoothing. Circumstantial changes could confound temporal dynamics. For instance, COVID-19 lockdown in 2020 reduced availability of treatment and prevention resources, but also reduced social activity and testing, altogether resulting in elevated growth of some PWID clusters.^49^ Although temporal trends were robust to serial intervals, R_e_ estimates remain uncertain due to noise, diagnosis delay, and incomplete sampling.

PrEP has contributed to reducing HIV transmission in BC, alongside interventions such as combination ART,^50,51^ TaSP,^52,53^ and safe injection sites.^54^ Ongoing evaluations of the effectiveness of interventions including PrEP access, education, and delivery modes in clusters and key populations contributes towards evidence for intervention adoption in other jurisdictions, and to improve the efficiency of treatment and prevention resources locally. Our findings highlight the success of layering PrEP aimed to individuals at high risk of HIV infection on top of generalized free access to ART, under the BC TasP strategy, to optimize the control of HIV/AIDS. At the same time, longitudinal phylogenetic monitoring offers the unique advantage of identifying individuals and groups that could benefit from prioritized ART and prevention education, access, and support.

## Contributors

AM and JBJ conceived of the study and AM designed the study. JT, PS, and CJB facilitated the data request. AM conducted analyses, generated figures, and wrote the first version of the draft. JT and CJB provided methodological feedback. JT, VM, PS, JT, MH, CJB, RB, JSGM, and JBJ provided edits. JBJ, JSGM, and AM received funding for the study. AM and JBJ had access to data.

## Data sharing

The BC-CfE is prohibited from making individuals’ data publicly available due to provisions in our service contracts, institutional policy, and ethical requirements. Code available upon request.

## Acknowledgements

We thank participants in the Drug Treatment Program and colleagues in the BC-CfE Molecular Laboratory and Drug Treatment Program. AM was supported by a doctoral grant from the Canadian Institutes of Health Research and the BC-CfE. JSGM is supported by grants paid to his institution by the British Columbia Ministry of Health, Health Canada, Public Health Agency of Canada, Vancouver Coastal Health, Vancouver General Hospital Foundation, and Genome British Columbia. JBJ is supported by the Canadian Institutes of Health Research, Genome British Columbia, Genome Canada, the Public Health Agency of Canada, and the BC-CfE. VM, PS, JT, MH, CJB, and RB were all supported by the BC-CfE.

