## Supplementary Material for "Impact of pre-exposure prophylaxis on HIV-1 drug resistance and phylogenetic cluster growth in British Columbia, Canada"

#### Table of Contents

Supplementary Methods ... 2

- Sequence cleaning, alignment, and subtype assignment ... 2
- PrEP eligibility and regimens in BC ... 2
- PrEP dispensation patterns of newly diagnosed PrEP users ... 3
- ART regimens of newly diagnosed PrEP users ... 4
- Viral lineage-level diversification rate ... 5
- Estimating HIV cases averted in phylogenetic clusters with stochastic branching processes ... 5

Supplementary Results ... 8

- Sociodemographic and clustering characteristics of newly diagnosed with and without PrEP ... 8
- Phylogenetic clustering of newly diagnosed individuals with and without PrEP ... 11
- Comparison of cluster identification under different tree rooting strategies ... 13
- Baseline drug resistance of newly diagnosed individuals with and without PrEP ... 14
- Drug resistance scores of newly diagnosed individuals with and without PrEP ... 17
- Active HIV-1 phylogenetic clusters in BC ... 18
- HIV effective reproduction number in British Columbia and key populations ... 27
- Phylogenetic cluster-specific effective reproduction number ... 35
- Counterfactual modeling of cluster growth ... 41

References ... 45

### Supplementary Methods

#### Sequence cleaning, alignment, and subtype assignment

There were 42,043 HIV-1 partial pol (*protease* and partial *reverse transcriptase*) sequences from 10,740 DTP participants collected between May 30, 1996 and December 31, 2022. Sequences were removed if they had no patient identifier (n=17) or duplicate laboratory ENUM (n=101), leaving 41,941 pol sequences (1-50 sequences per patient). For 20 sequences, collection date was imputed as the test request date plus the median difference between collection date and request date for all other samples in that year. Data included 8759 *integrase* (int) sequences collected August 21, 1996 to December 31, 2022 representing 4058 unique patients. There were 309 int sequences without a collection date, for which we queried the sample collection date from the matching sample in the pol sequence; 20 sequences were removed because no matching sample date was available leaving 8739 sequences from 4052 patients (1-35 sequences per patient). Partial pol and int sequences were aligned to the HXB2 reference genome (GenBank Accession #K03455) using minimap2 parallelized by viralMSA.[1,2] COMET was used to assign HIV-1 subtypes for partial pol.[3] For COMET subtype assignments with less than 90% bootstrap support, subtype assignment was evaluated using REGA,[4] and the assignment with higher support was chosen. 41,477 subtypes were assigned with COMET and 95 with REGA. Codons of surveillance drug resistance mutations (SDRMs)[5] and insertions relative to reference HXB2 were removed prior to phylogenetic inference.

#### PrEP eligibility and regimens in BC

As per guidance for the use of PrEP in BC updated in 2020, individuals must be at substantial risk of acquiring HIV and clinically eligible (HIV negative test within 15 days, no signs of HIV infection, normal renal function, and hepatitis B virus infection status).[6] For people who inject drugs (PWID), that constitutes reporting shared injection equipment with a HIV virally unsuppressed injecting partner. Heterosexual (HET) men and women are eligible if they report condomless intercourse with a virally unsuppressed partner. For gay, bisexual, and other men who have sex with men (GBM), eligibility is determined based on reporting condomless sex and a recent syphilis, gonorrhea, or chlamydia infection; an ongoing sexual relationship with an HIV-positive partner with detectable viral load; repeated courses of post-exposure prophylaxis; or a HIV Incidence Risk Index (HIRI) score greater than or equal to 10. A HIRI score is calculated on the basis of age, number of male sex partners, frequency of receptive anal intercourse, number of HIV positive male sex partners, frequency of unprotected anal sex with someone who is HIV positive, and recent use of methamphetamines.[6]

Truvada (emtricitabine/tenofovir disoproxil fumarate; FTC/TDF) is an oral PrEP regimen recommended for daily use, but an on-demand dosing schedule may be considered for cis-gender MSM, following the “2-

1-1" dosage with 2 pills 2-24 hr before sex and 1 pill daily until 48 hours after sex.[7] Individuals prescribed PrEP are required to return for follow-up visits after 1 month and at minimum every 3 months for HIV and STI testing, renal function analysis, adherence counselling, risk reduction support, side effect assessment, and STI symptom assessment. In this study, we did not differentiate between daily versus on-demand PrEP usage. Other PrEP delivery systems including oral tenofovir alafenamide, and emtricitabine (FTC/TAF, Descovy; approved by Health Canada in December 2020, but not commonly prescribed in BC unless patient has renal or bone dysfunction), long-acting injectable cabotegravir (CAB-LA; first CAB-LA PrEP, Apretude, approved in Canada in May 2024)[8], dapivirine vaginal rings (licensed in several African nations), and lenacapavir, are not included in this study.

#### PrEP dispensation patterns of newly diagnosed PrEP users

We compared PrEP refill characteristics of newly diagnosed PrEP users with and without baseline drug resistant mutations. PrEP adherence more broadly in BC has been reported previously.[9,10] Using data on PrEP requisition date among newly diagnosed with PrEP use, dispensation date, and doses over time, we quantified the number of prescriptions filled; total dosage days; time from last prescription dispensation to date of first detectable viral load; time from last prescription dispensation to first antiretroviral date; for those with multiple prescriptions, the total, mean, median, minimum, and maximum days with no refill (from last dosage day to next prescription filled) and the percentage of days with no refill (1 – proportion of days covered).

Forty-two individuals were prescribed PrEP and later diagnosed with HIV; however, three never filled their prescription, leaving 39 newly diagnosed PrEP users. Newly diagnosed PrEP users had one to 14 PrEP prescriptions filled (median=2, mean=3.0), and 25 individuals had more than one prescription. Total dosage days ranged from 30 d to 1230 d (median=120 d; mean=219.4 d). The time from last PrEP dispensation to date of first detectable viral load ranged widely from -4 d (PrEP was likely prescribed before requisite HIV test result available) to 1605 d (median=331 d; mean=494.0 d). The distribution of times from last PrEP dispensation to date of first detectable viral load was not significantly different between those with and without baseline NRTI resistance (medians=54 d vs. 343 d; Kruskal-Wallis test, p=0.065). Time from last PrEP dispensation to date of first antiretroviral ranged from 8 d to 1611 d (median=364 d; mean=496.8 d). Those with baseline NRTI resistance had significantly less time from last PrEP dispensation to date of first detectable viral load (medians=76 vs. 373 d; Kruskal-Wallis test, p=0.045). Six individuals had recent PrEP use, similar to the definition of Misra et al.,[11] with time from most recent dispensation to estimated seroconversion less than or equal to 90 days; two of six individuals with recent PrEP use (29 and 54 days from last dispensation to seroconversion) had baseline NRTI resistance. Nine individuals had less than 90 days from the end of their last prescribed dosage date to first detectable viral load.

For PrEP users with multiple prescriptions filled (n=20), the total number of days with no refill (from last dosage day to next prescription filled, across all prescriptions) ranged from -15 d – 753 d (median=36.5 d; mean=121.2 d), and was not significantly different between those with and without baseline NRTI resistance (Kruskal test: p=0.11). Nor were there significant differences between the median, maximum, or proportion of days with no refill between these groups (Kruskal test: p=0.36; p=0.052; p=0.052). Two PrEP users with baseline NRTI resistance who had multiple prescriptions filled also had the second and fourth highest proportion of days with no refill (0.037 and 0.40), as well as the fourth and fifth highest total number of days with no refill (203 and 290). This could be indicative of on-demand PrEP use, smaller than recommended dosing, or incomplete adherence.

#### ART regimens of newly diagnosed PrEP users

We considered the frequencies of commonly prescribed NRTI drugs in first and most recent antiretroviral therapy (ART) regimens among newly diagnosed PrEP users. The majority of PrEP users were initially prescribed an ART regimen containing TDF (n=15) and/or FTC (n=38). Of their most recent drug regimens, 33 were still prescribed FTC and one was still prescribed TDF (**Table S2**).

**Table S1. HIV-1 antiretroviral therapy drug names, codes, and classes.** Drug classes include NRTI = nucleoside-analogue reverse transcriptase inhibitor; NNRTI = non-nucleoside-analogue reverse transcriptase inhibitor; PI = protease inhibitor; and INSTI = integrase strand inhibitor. Excludes broadly neutralizing antibodies, capsid inhibitors, other classes). Components of TDF/FTC oral PrEP in **bold**.

| **Name** | **Code** | **Class** |
|---|---|---|
| **Emtricitabine** | **FTC** | **NRTI** |
| **Tenofovir disoproxil fumarate** | **TDF** | **NRTI** |
| Tenofovir alafenamide fumarate | TAF | NRTI |
| Lamivudine | 3TC | NRTI |
| Abacavir | ABA/ABC | NRTI |
| Zidovudine | AZT | NRTI |
| Stavudine | D4T | NRTI |
| Zalcitabine | DDC | NRTI |
| Didanosine | DDI | NRTI |
| Efavirenz | DMP | NNRTI |
| Etravirine | ETV | NNRTI |
| Nevirapine | NEV | NNRTI |
| Rilpivirine | RPV | NNRTI |
| Atazanavir | ATA | PI |
| Darunavir | DRV | PI |
| Indinavir | IND | PI |
| Nelfinavir | NEL | PI |
| Saquinavir | SAQ | PI |
| Tipranavir | TIP | PI |
| Bictegravir | BCG | INSTI |
| Dolutegravir | DTG | INSTI |
| Elvitegravir | EGV | INSTI |
| Raltegravir | MKS | INSTI |

**Table S2. First and most recent drug regimens of newly diagnosed PrEP users.**

| Drug regimen | First regimen (n) | Most recent regimen (n) |
|---|---|---|
| **FTC**/TAF/BCG/ | 22 | 32 |
| **TDF/FTC**/DRV/DTG/COB/ | 12 | 0 |
| 3TC/ABA/DTG/ | 0 | 4 |
| **TDF/FTC**/DTG/ | 2 | 0 |
| 3TC/DTG/ | 0 | 2 |
| 3TC/ABA/DRV/COB/ | 1 | 0 |
| **FTC**/DRV/COB/TAF/BCG/ | 1 | 0 |
| **TDF/FTC**/DRV/COB/ | 1 | 0 |
| **TDF/FTC**/DTG/ | 0 | 1 |

#### Lineage-level viral diversification rate

In addition to comparing frequencies of clustering, we also evaluated whether newly diagnosed PrEP users and non-PrEP users differed in their lineage-level viral diversification rates, a metric reflecting historical lineage branching frequency, calculated using trees pruned to the oldest sequence per patient. Lineage-level viral diversification rate for each tip on a rooted bifurcating tree is the reciprocal sum of $N_i$ branch lengths ($l_j$) from tip $i$ to the root, with each consecutive edge ($j$) down-weighted by a factor of 1⁄2.[12] For each tip, the mean lineage-level diversification rate across 100 bootstrap trees was computed.

$$Lineage - level\ viral\ diversification\ rate_i = (\sum_{j=1}^{N_i} \frac{l_j}{2^{j-1}})^{-1}$$

#### Estimating HIV cases averted in phylogenetic clusters with stochastic branching processes

We applied stochastic branching processes to simulate HIV cluster growth with and without widespread PrEP availability, where the number of secondary cases follows a negative binomial distribution defined by mean $R_e$ and dispersion k,[13,14] reflecting infrequent but characteristic superspreading events.[15] We assumed a gamma-distributed serial interval with mean 1 y and sd 0.5 y; k sampled uniformly between 0.1 and 0.3; and $R_e$ specified as observed cluster $R_e$ (with PrEP available) on that date or as cluster $R_e$ adjusted by a PrEP effect, which is the FC $R_e$ with PrEP (**Fig. 3C, Fig. S21, Fig. S22**). Observed cluster $R_e$ with 95% confidence width greater than 10 (sparse data) or NA were replaced with $R_e$=0.8 to reflect inactivity. If PrEP effect could not be calculated for a cluster, either because it was a new cluster seeded since 2018 or Re was NA before or after PrEP, then the PrEP effect of the clusters' predominant population was assigned. For each generation, $R_e$ was permitted to deviate from the specified value by drawing from a binomial distribution with 50% probability of success, where $R_e$ was then drawn from a gamma-distribution centered on cluster $R_e$ (sd=0.1).

The starting seed (number of infectious individuals on January 1, 2018) for each cluster was calibrated to align the observed number of new samples in each cluster from 2018 to end of 2022, with the mean number of sampled new cases from simulations using observed cluster $R_e$ (**Table S3**, **Fig. S23**). Initially, the number of infectious individuals at time zero was specified as cumulative cluster cases minus deaths and emigrations at the end of 2017 multiplied by the proportion of diagnosed cases on ART and not virally suppressed, based on estimates of progress towards 95-95-95 goals in BC from 2019, in which 82% of cases in BC were diagnosed, 76% of those diagnosed were on ART, and 83% of those on ART were virally suppressed.[16] The adjusted seed size was applied in all subsequent simulations based on observed $R_e$ (with PrEP available) and counterfactual $R_e$ (in the absence of PrEP). Seeds were enforced to be a minimum of one, including for clusters seeded since 2018 (e.g., cluster 114).

For scenarios with or without PrEP, 4000 simulations were run per cluster and for non-clustered cases (grouped as cluster 9999). Infected cases resulting from simulations were stochastically diagnosed with probability sampled from a log-normal distribution centered on 0.82, and diagnosed cases were sampled (i.e., sequenced) with probability drawn from a log-normal distribution centered on 0.95 (informed by ratio of DTP participants since 2018 with a pol sequence). Simulations began on day zero, January 1, 2018 and ran to January 1, 2023 (1827 d). New and cumulative diagnoses and samples over time were calculated. Bootstrapping was used to calculate 95% confidence intervals of diagnoses or samples averted by sampling 95% of simulations without removal 1000 times, then calculating normally-distributed confidence intervals across bootstrap means. Cluster diagnoses averted were calculated as the difference in mean and 95% CI of cumulative diagnoses in simulations with and without PrEP (**Fig. S24, Fig. S25**). Poisson generalized linear models were evaluated to identify cluster characteristics associated with the most and fewest diagnoses averted (**Table S9**). Diagnoses or samples averted were shifted up by the minimum averted to remove negative integers; factors evaluated included cluster size in 2017, predominant and percentage key populations, percentage health authority of residence, median member age in 2023 and age at first ART, and percentage of new diagnoses with PrEP experience.

**Table S3. Estimation of simulation cluster seed size (number of infectious individuals at time zero).** Active clusters shown. Cluster 9999 represents non-clustered individuals.

| Cluster ID | Initial seed | N new samples observed | Mean simulated N new samples (observed $R_e$) | Adjust factor | Adjusted seed |
|---|---|---|---|---|---|
| 9999 | 116 | 439 | 205 | 0.4669 | 249 |
| 57 | 17 | 19 | 8 | 0.4210 | 41 |
| 31 | 5 | 94 | 16 | 0.1702 | 30 |
| 49 | 17 | 11 | 18 | 1.6363 | 11 |
| 202 | 1 | 9 | 1 | 0.1111 | 10 |
| 168 | 1 | 17 | 2 | 0.1176 | 9 |
| 207 | 1 | 6 | 1 | 0.1667 | 6 |
| 197 | 1 | 5 | 1 | 0.2 | 5 |
| 40 | 1 | 3 | 1 | 0.3333 | 4 |
| 52 | 1 | 4 | 1 | 0.25 | 4 |
| 134 | 1 | 7 | 2 | 0.2857 | 4 |
| 196 | 1 | 3 | 1 | 0.3333 | 4 |
| 95 | 2 | 27 | 18 | 0.6667 | 3 |
| 137 | 3 | 26 | 28 | 1.0769 | 3 |
| 13 | 3 | 3 | 8 | 2.6667 | 2 |
| 26 | 1 | 5 | 4 | 0.8 | 2 |
| 142 | 2 | 6 | 8 | 1.3333 | 2 |
| 182 | 1 | 3 | 2 | 0.6667 | 2 |
| 201 | 3 | 2 | 4 | 2 | 2 |
| 209 | 1 | 5 | 3 | 0.6 | 2 |
| 217 | 1 | 4 | 3 | 0.75 | 2 |
| 219 | 1 | 4 | 2 | 0.5 | 2 |
| 234 | 1 | 6 | 4 | 0.6667 | 2 |
| 194 | 1 | 5 | 4 | 0.8 | 2 |
| 114 | 1 | 14 | 9 | 0.6428 | 2 |
| 205 | 1 | 9 | 5 | 0.5555 | 2 |
| 14 | 1 | 1 | 2 | 2 | 1 |
| 22 | 1 | 19 | 24 | 1.2631 | 1 |
| 29 | 2 | 1 | 3 | 3 | 1 |
| 33 | 1 | 1 | 4 | 4 | 1 |
| 35 | 1 | 2 | 2 | 1 | 1 |
| 41 | 1 | 7 | 10 | 1.4285 | 1 |

### Supplementary Results

#### Sociodemographic and clustering characteristics of newly diagnosed with and without PrEP

We investigated differences in sociodemographic and clinical factors associated with newly diagnosed with or without PrEP use (**Table S4**) and associated with cluster membership stratified by newly diagnosed individuals with PrEP use or not (**Table S5**). Among PrEP users, lower baseline CD4 counts were associated with clustering (Kruskal test, p=0·011), whereas in non-PrEP users, higher baseline CD4 counts were associated with clustering (p<0·001). Among newly diagnosed non-PrEP users, other factors associated with elevated risk of clustering included being a person who injects drugs (PWID; p<0·001), having previous HCV infection (p<0·001), which is colinear with PWID, and living in Vancouver Island health authority (p<0·001). Newly diagnosed non-PrEP users were less likely to cluster if they had heterosexual risk exposure (p=0·0015), resided in Interior or Fraser health authorities (p=0·014, p<0·001), or were infected with subtypes C, 01_AE, A1, or A6 (all p<=0·001). Since 2018, there have been new cluster members with subtypes F1 (n=2), 01_AE (n=1), B recombinant (n=1), and a B, D recombinant (n=1), however these assignments have relatively low bootstrap support and occurred in clusters predominantly comprised of subtype B (**Table S5**).

**Table S4. Characteristics of newly diagnosed PrEP users and non-PrEP users from 2018-2022.** Proportions of each parameter compared using chi-squared test and medians compared using Kruskal-Wallis test.

| | | Previous PrEP users | | Non-PrEP users | | |
|---|---|---|---|---|---|---|
| | **Parameter** | **n** | **%** | **n** | **%** | **p-value** |
| | **Total** | 39 | 100 | 566 | 100 | - |
| **Gender** | Female | 0 | 0 | 83 | 14.7 | **1.60E-02*** |
| | Male | 39 | 100 | 450 | 79.5 | **1.09E-02*** |
| | Trans Male | 0 | 0 | 0 | 0 | - |
| | Trans Female | 0 | 0 | 6 | 1.1 | 1 |
| **Risk Exposure** | Not reported | 4 | 10.3 | 99 | 17.5 | 3.46E-01 |
| | GBM | 34 | 97.1 | 265 | 56.7 | **9.57E-06*** |
| | HET | 1 | 2.9 | 152 | 32.5 | **7.76E-04*** |
| | PWID | 3 | 8.6 | 114 | 24.4 | 7.99E-02 |
| | HCV | 2 | 5.1 | 109 | 19.3 | 5.08E-02 |
| | HBV | 0 | 0 | 36 | 6.4 | **6.46E-03*** |
| **Health Authority** | Not reported | 0 | 0 | 26 | 4.6 | 3.16E-01 |
| | Interior | 1 | 2.6 | 36 | 6.7 | 5.01E-01 |
| | Fraser | 11 | 28.2 | 139 | 25.7 | 8.81E-01 |
| | Vancouver Coastal | 24 | 61.5 | 241 | 44.6 | 6.00E-02 |
| | Vancouver Island | 3 | 7.7 | 71 | 13.1 | 4.61E-01 |
| | Northern | 0 | 0 | 27 | 5 | 3.00E-01 |
| **Subtype** | B | 37 | 94.8 | 465 | 82.2 | 6.82E-02 |
| | C | 0 | 0 | **39** | 6.9 | 1.75E-01 |
| | 01_AE | 0 | 0 | **24** | 4.2 | 3.74E-01 |
| | A1 | **1** | 2.6 | **11** | 1.9 | 1 |
| | A6 | 0 | 0 | **7** | 1.6 | 1 |
| | 12_BF | 0 | 0 | **3** | 0.53 | 1 |
| | G | 0 | 0 | **3** | 0.53 | 1 |
| | D | 0 | 0 | **2** | 0.35 | 1 |
| | F1 | 0 | 0 | **2** | 0.35 | 1 |
| | 02_AG | **1** | 2.6 | **1** | 0.18 | 2.85E-01 |
| | 06_cpx | 0 | 0 | **1** | 0.18 | 1 |
| | 07_BC | 0 | 0 | **1** | 0.18 | 1 |
| | 33_01B | 0 | 0 | **1** | 0.18 | 1 |
| | 35_A1D | 0 | 0 | **1** | 0.18 | 1 |
| | 44_BF1 | 0 | 0 | **1** | 0.18 | 1 |
| | A3 | 0 | 0 | **1** | 0.18 | 1 |
| | B recombinant | 0 | 0 | **1** | 0.18 | 1 |
| | CRF 19_cpx | 0 | 0 | **1** | 0.18 | 1 |
| | B, D recombinant | 0 | 0 | **1** | 0.18 | 1 |
| | | **Median** | **-** | **Median** | - | |
| | log(pVL) | 4.76 | - | 4.8 | - | 6.51E-01 |
| | CD4 baseline | 490 | - | 380 | - | **3.45E-03*** |
| | Age at first ARV | 32 | - | 37 | - | **3.26E-02*** |

**Table S5. Sociodemographic and clinical factors associated with clustering among newly diagnosed PrEP users and non-PrEP users from 2018-2022.** P-values reported for univariate two-sided chi-squared tests for categorical variables and Kruskal-Wallis tests for numeric variables.

| | | Newly diagnosed PrEP users | | | | Newly diagnosed non-PrEP users | | | |
|---|---|---|---|---|---|---|---|---|---|
| | **Parameter** | **Total** | **n clust.** | **% clust.** | **p-value** | **Total** | **n clust.** | **% clust.** | **p-value** |
| | **n** | 39 | 30 | 76.9 | - | 566 | 303 | 53.5 | - |
| **Gender** | **Female** | 0 | 0 | - | - | 83 | 40 | 48.2 | 2.61E-01 |
| | **Male** | 39 | 30 | 76.9 | - | 450 | 251 | 55.8 | 2.02E-01 |
| | **Trans Male** | 0 | 0 | - | - | 0 | 0 | - | - |
| | **Trans Female** | 0 | 0 | - | - | 6 | 3 | 50.0 | 1 |
| **Risk Exposure** | **Not reported** | 4 | 2 | 50.0 | 4.70E-01 | 99 | 43 | 43.4 | 1.85E-01 |
| | **GBM** | 34 | 28 | 82.4 | 6.29E-02 | 265 | 145 | 54.7 | 3.11E-01 |
| | **HET** | 1 | 0 | 0.0 | 6.29E-02 | 152 | 68 | 44.7 | **1.52E-03** |
| | **PWID** | 3 | 3 | 100.0 | 2.78E-01 | 114 | 91 | 79.8 | **6.94E-09** |
| | **HCV** | 2 | 2 | 100.0 | 7.28E-01 | 109 | 86 | 78.9 | **6.98E-08** |
| | **HBV** | 0 | 0 | - | 8.25E-01 | 36 | 14 | 38.9 | 1.45E-01 |
| **Health Authority** | **Not reported** | 0 | 0 | - | - | 26 | 9 | 34.6 | 8.89E-01 |
| | **Interior** | 1 | 1 | 100.0 | 1 | 36 | 12 | 33.3 | **1.39E-02** |
| | **Fraser** | 11 | 9 | 81.8 | 9.74E-01 | 139 | 56 | 40.3 | **1.50E-04** |
| | **Van. Coastal** | 24 | 17 | 70.8 | 4.53E-01 | 241 | 136 | 56.4 | 4.56E-01 |
| | **Van. Island** | 3 | 3 | 100.0 | 7.84E-01 | 71 | 55 | 77.5 | **5.09E-05** |
| | **Northern** | 0 | 0 | - | - | 27 | 20 | 74.1 | 5.70E-02 |
| **Subtype** | **B** | 37 | 30 | 81.1 | 7.36E-02 | 465 | 298 | 64.1 | **1.13E-26** |
| | **C** | 0 | 0 | - | - | 39 | 0 | 0.0 | **1.20E-11** |
| | **01_AE** | 0 | 0 | - | - | 24 | 1 | 4.2 | **2.07E-06** |
| | **A1** | 1 | 0 | 0.0 | 5.17E-01 | 11 | 0 | 0.0 | **1.00E-03** |
| | **A6** | 0 | 0 | - | - | 7 | 0 | 0.0 | **1.00E-03** |
| | **12_BF** | 0 | 0 | - | - | 3 | 0 | 0.0 | 1.99E-01 |
| | **G** | 0 | 0 | - | - | 3 | 0 | 0.0 | 1.99E-01 |
| | **D** | 0 | 0 | - | - | 2 | 0 | 0.0 | 4.18E-01 |
| | **F1** | 0 | 0 | - | - | 2 | 2 | 100.0 | 5.42E-01 |
| | **02_AG** | 1 | 0 | 0.0 | 5.17E-01 | 1 | 0 | 0.0 | 9.43E-01 |
| | **06_cpx** | 0 | 0 | - | - | 1 | 0 | 0.0 | 9.43E-01 |
| | **07_BC** | 0 | 0 | - | - | 1 | 0 | 0.0 | 9.43E-01 |
| | **33_01B** | 0 | 0 | - | - | 1 | 0 | 0.0 | 9.43E-01 |
| | **35_A1D** | 0 | 0 | - | - | 1 | 0 | 0.0 | 9.43E-01 |
| | **44_BF1** | 0 | 0 | - | - | 1 | 0 | 0.0 | 9.43E-01 |
| | **A3** | 0 | 0 | - | - | 1 | 0 | 0.0 | 9.43E-01 |
| | **B recomb.** | 0 | 0 | - | - | 1 | 1 | 100.0 | 1 |
| | **CRF 19_cpx** | 0 | 0 | - | - | 1 | 0 | 0.0 | 9.43E-01 |
| | **B, D recomb.** | 0 | 0 | - | - | 1 | 1 | 100.0 | 1 |
| | | **Median, all** | **Median, clust.** | | | **Median, all** | **Median, clust.** | | |
| | **log10 (plasmaVL)** | 4.76 | 4.75 | - | 9.47E-01 | 4.8 | 4.85 | - | 1.12E-01 |
| | **CD4 baseline** | 490 | 407 | - | **1.13E-02** | 380 | 440 | - | **8.09E-05** |
| | **Age first ARV** | 32 | 32.5 | - | 7.39E-01 | 37 | 36 | - | 2.57E-01 |

#### Phylogenetic clustering of newly diagnosed individuals with and without PrEP

**Table S6. Clustering among newly diagnosed overall from 2015-2022, and since PrEP availability in 2018, stratified by PrEP use**. Sum/mean clustering across 2018-2022 reported on bottom row.

| | Overall newly diagnosed | | | | Newly diagnosed, no PrEP | | | | Newly diagnosed, PrEP | | | |
|---|---|---|---|---|---|---|---|---|---|---|---|---|
| **Year** | **N** | **N clustering** | **% clustering** | **N unique clusters** | **N** | **N clustering** | **% clustering** | **N unique clusters** | **N** | **N clustering** | **% clustering** | **N unique clusters** |
| 2015 | 256 | 140 | 54.7 | 48 | | | | | | | | |
| 2016 | 252 | 121 | 48 | 45 | | | | | | | | |
| 2017 | 183 | 103 | 56.3 | 43 | | | | | | | | |
| 2018 | 174 | 94 | 54 | 40 | 173 | 93 | 53.8 | 40 | 1 | 1 | 100 | 1 |
| 2019 | 168 | 83 | 49.4 | 43 | 160 | 78 | 48.8 | 42 | 8 | 5 | 62.5 | 4 |
| 2020 | 127 | 83 | 65.4 | 30 | 121 | 78 | 64.5 | 30 | 6 | 5 | 83.3 | 5 |
| 2021 | 131 | 74 | 56.5 | 34 | 125 | 70 | 56 | 34 | 6 | 4 | 66.7 | 3 |
| 2022 | 124 | 67 | 54 | 24 | 106 | 52 | 49.1 | 24 | 18 | 15 | 83.3 | 7 |
| **Sum/Mean: 2018-2022** | | | | | 685 | 371 | 54.2 | 84 | 39 | 30 | 76.9 | 13 |

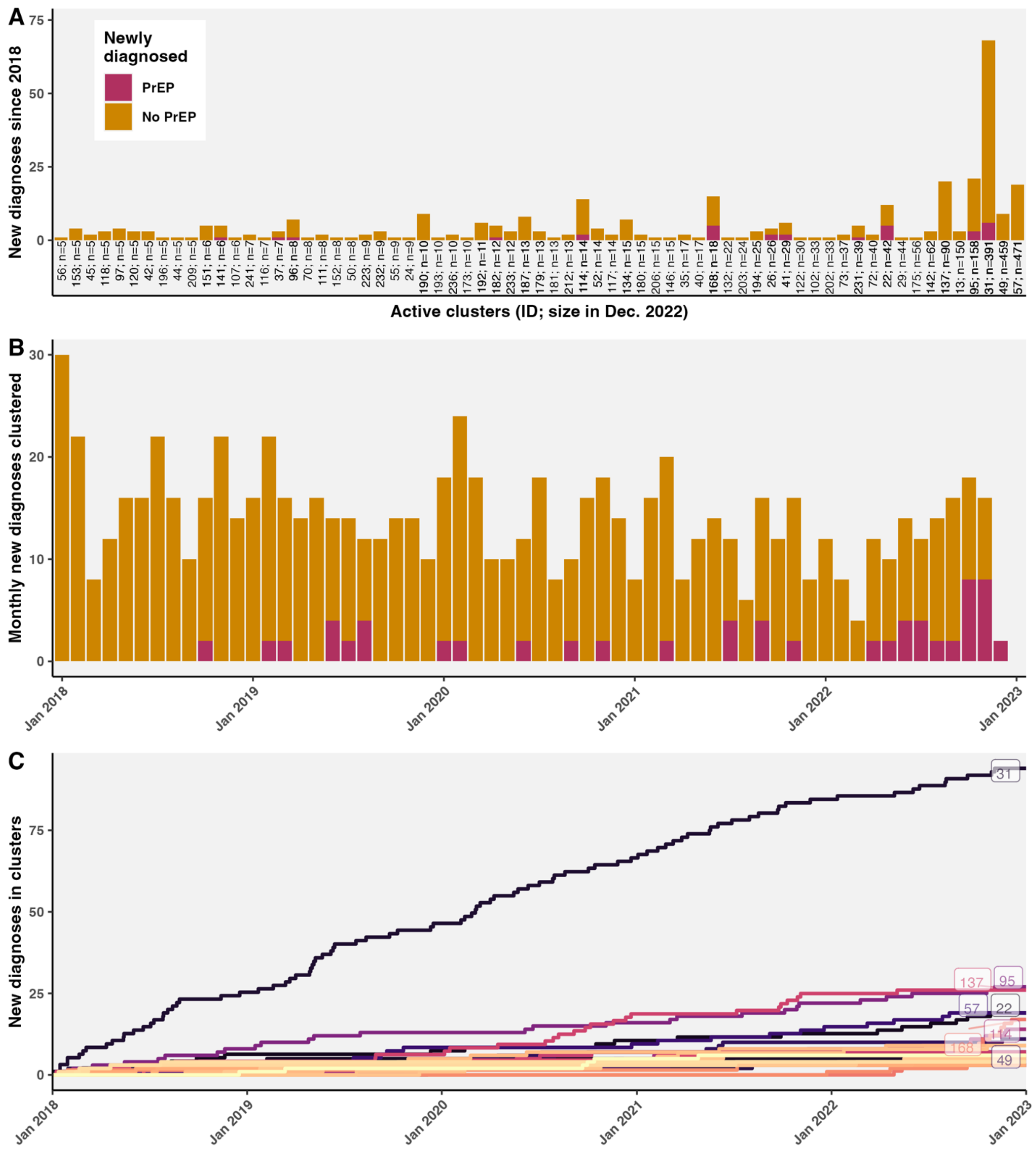


**Fig. S1. New diagnoses in active phylogenetic clusters since 2018.** A) Total new diagnoses in active clusters, annotated with cluster ID; cluster size at the end of study period in December 2022. Bars are colored by PrEP use. B) Monthly new diagnoses that clustered during the study period, colored by PrEP use. C) New diagnoses in the study period by cluster. Annotated with cluster ID for the eight largest clusters.

In corroboration of elevated clustering, PrEP users had significantly higher viral lineage-level diversification rates than non-PrEP users (**Fig. S2**; Mann-Whitney test, $p<0{\cdot}001$). Viral lineage-level diversification rates were not significantly different between PrEP users and non-PrEP users who clustered (Mann-Whitney test: p=0·16) or did not cluster (p=0·50).

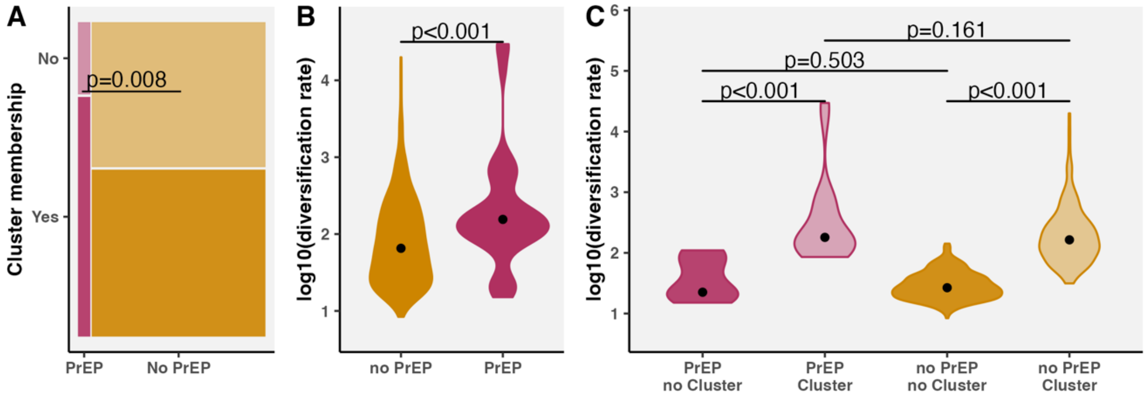


**Fig. S2. Clustering and lineage-level viral diversification rate among newly diagnosed with and without PrEP use. A)** Comparison of the proportion of individuals who clustered (membership, yes or no) by PrEP use, using a two-sided chi-squared test. **B)** Distribution of viral lineage-level diversification rate among newly diagnosed PrEP users and non-PrEP users, compared using a Kruskal-Wallis test**. C)** Viral lineage-level diversification rates were compared by PrEP use and clustering using pairwise Mann-Whitney tests.

#### Comparison of cluster identification under different tree rooting strategies

We investigated the sensitivity of cluster identification to rooting strategies for maximum likelihood divergence-scaled HIV-1 phylogenetic trees. Starting with the same set of 100 bootstrap partial pol trees from FastTree, we compared the default midpoint-rooted (MPR) trees, a binarized MPR tree, to those rooted using three different outgroups (oldest subtype B, G, and H in the BC data), along with a subset of 10 trees, due to computational feasibility, rooted under the assumption of a strict molecular clock using root-to-tip regression in TempEst,[17] and we compared clusters identified using a 50, 70, and 90% threshold for the

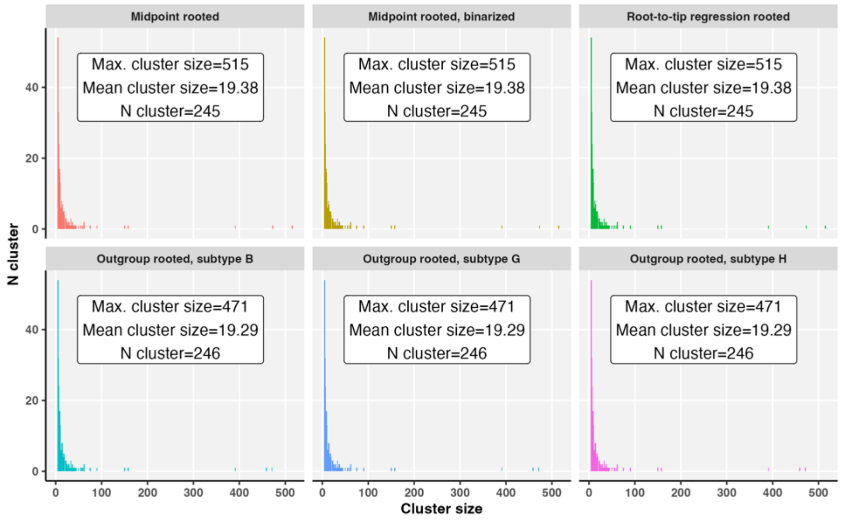

percent of bootstrap trees connecting individuals. We identified clusters with 5 or more members linked by less than 0.02 substitutions/site tree distance, and then compared the number of clusters and distribution of cluster size (**Fig. S3**).

**Fig. S3. Distribution of phylogenetic cluster sizes identified using multiple rooting strategies.** Trees were rooted using midpoint (MPR; Fasttree default), MPR binarized, root-to-tip regression, or outgroup rooting on oldest subtype B, G, or H. Annotations for the maximum and mean cluster size, and total number of clusters.

#### Baseline drug resistance of newly diagnosed individuals with and without PrEP

Of NRTI resistance-associated mutations (RAMs), NRTI K70R was most common at baseline in the entire cohort and is increasing in prevalence over time (**Fig. S4-6**). The next most frequently detected baseline mutations were NRTI resistance mutation, K103N, and NNRTI resistance mutation, T215E. K103N has been identified independently in six active (defined as new cases since 2018) phylogenetic clusters, while baseline T215E is present in all 14 members of cluster 114 (**Fig. S13**).

**Table S7. Proportion with treatment selected mutations among newly diagnosed PrEP users and non-PrEP users from 2018-2022.** Frequencies were compared using Fisher's exact tests (as the assumption of minimum expected frequencies for chi-squared violated) and p-values were adjusted using Benjamini-Hochberg method. M184V including and excluding M184IV mixture are emboldened.

| | | | Newly Diagnosed PrEP users (n=39) | | Newly Diagnosed Non-PrEP users (n=566) | | Fisher's Test | |
|---|---|---|---|---|---|---|---|---|
| **Drug class** | **Mutation** | **Gene** | **Total** | **%** | **Total** | **%** | **Unadj. p-val** | **Adj. p-val** |
| **NRTI** | M41L | rt | 1 | 2.56 | 10 | 1.77 | 0.5226 | 0.5226 |
| | K70R | rt | 1 | 2.56 | 37 | 6.54 | 0.5012 | 0.5226 |
| | V75I | rt | 1 | 2.56 | 2 | 0.35 | 0.1815 | 0.3327 |
| | **M184V** | **rt** | **2** | **5.13** | **2** | **0.35** | **0.0224*** | **0.2463** |
| | **M184IV/V** | **rt** | **3** | **7.69** | **2** | **0.35** | **0.0023*** | **0.0250*** |
| | T215E | rt | 3 | 7.69 | 15 | 2.65 | 0.1031 | 0.2747 |
| **NNRTI** | K103N | rt | 2 | 5.13 | 15 | 2.65 | 0.3007 | 0.4725 |
| | K103S | rt | 1 | 2.56 | 8 | 1.41 | 0.4533 | 0.5226 |
| | Y181C | rt | 1 | 2.56 | 9 | 1.59 | 0.4891 | 0.5226 |
| | N348I | rt | 2 | 5.13 | 6 | 1.06 | 0.0886 | 0.2747 |
| **PI** | A71I | pro | 1 | 2.56 | 1 | 0.18 | 0.1249 | 0.2747 |
| | L90M | pro | 1 | 2.56 | 1 | 0.18 | 0.1249 | 0.2747 |

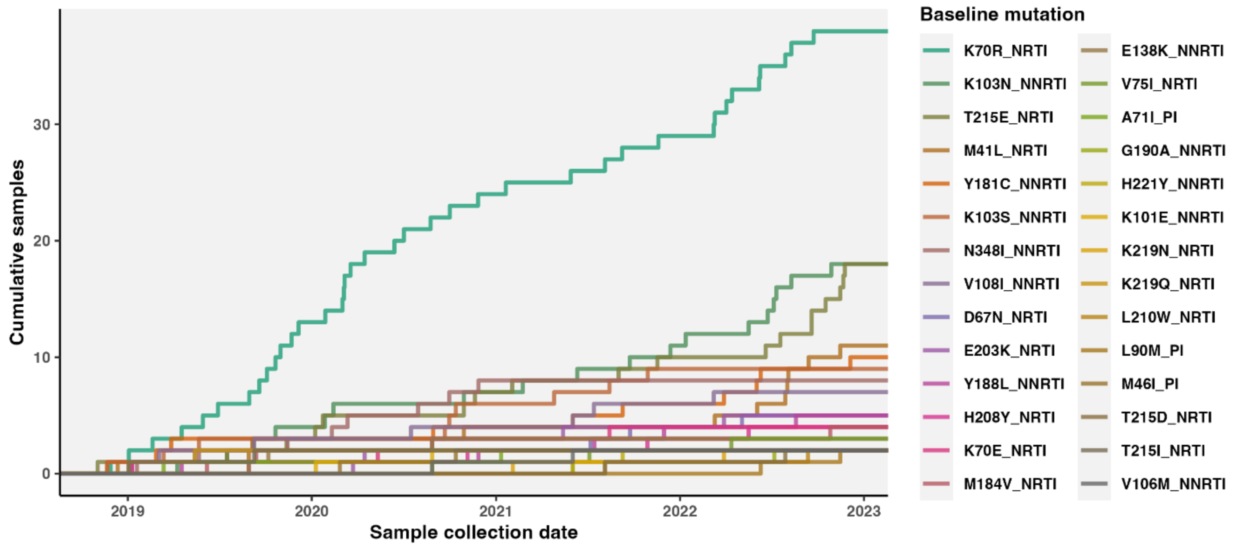


**Fig. S4. Cumulative detection of baseline treatment selected mutations among newly diagnosed 2018-2022.**

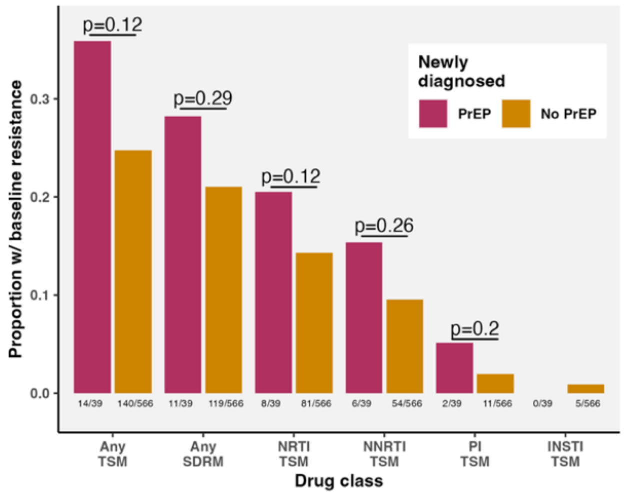


**Fig. S5. Proportion newly diagnosed with or without PrEP with any baseline drug resistance mutations by drug class.**

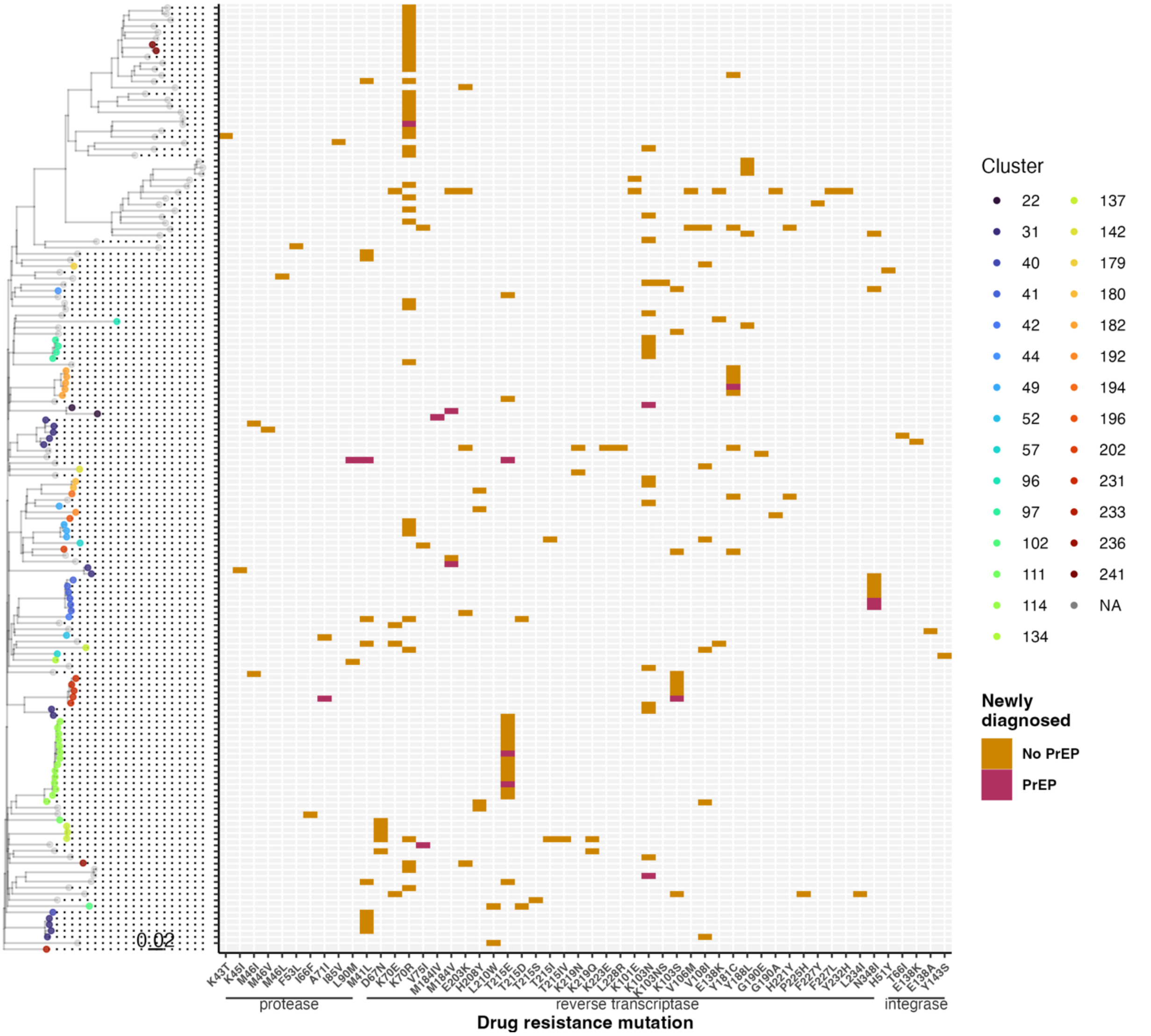


**Fig. S6. Relationships between baseline drug resistance mutations in newly diagnosed in BC with or without PrEP use.** Restricted to oldest sequences from participants with any baseline drug resistance diagnosed from 2018 to 2022 (n=154). Mutations sorted by gene, then codon position, and colored by PrEP use. Aligned phylogeny is a bootstrap approximate maximum likelihood tree of ~40,000 partial pol sequences trimmed of drug resistance mutation codons, rooted on the oldest subtype B sequence, and pruned to newly diagnosed with any baseline drug resistance mutations; scale in substitutions per site. Integrase DRMs were called from patient-matched integrase sequences not included in phylogeny.

**Drug resistance scores of newly diagnosed individuals with and without PrEP**

Differences in M184V manifested as elevated baseline drug resistance scores for lamivudine (3TC; Kruskal-Wallis, p<0·001), emtricitabine (FTC; p<0·0001), and abacavir (ABC; p=0·020) in PrEP users compared to non-PrEP users (**Fig. S5**). Newly diagnosed PrEP users also had significantly elevated didanosine (DDI) drug resistance scores (p=0·019), driven by co-occurring thymidine analog mutations, T215E and M41L.

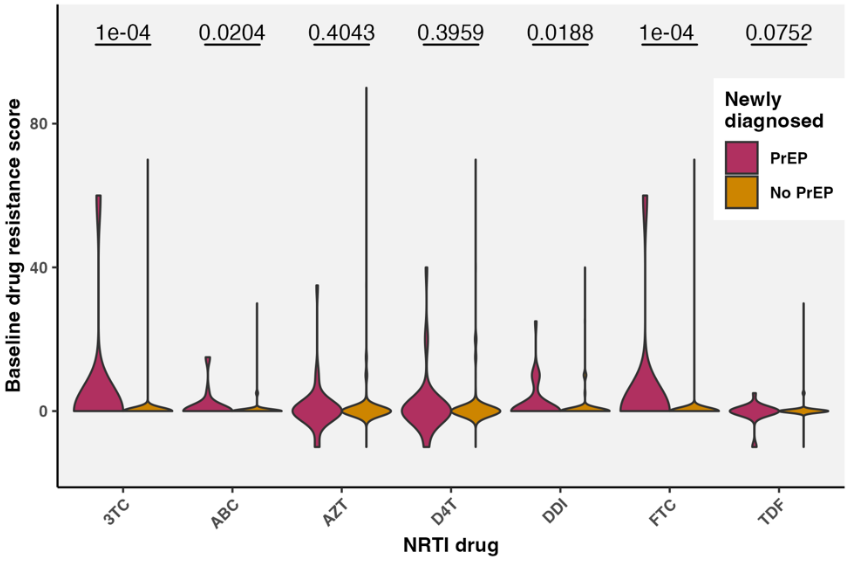


**Fig. S7. NRTI drug resistance scores using in newly diagnosed with and without PrEP**. Scores calculated using Stanford algorithm version 9.6 updated March 9, 2024. Drugs include 3TC (lamivudine), ABC (abacavir), AZT (zidovudine), D4T (stavudine), DDI (didanosine), FTC (emtricitabine), and TDF (tenofovir disoproxil fumarate).

#### Active HIV-1 phylogenetic clusters in BC

Active clusters with at least 1 new case since January 1, 2018 were characterized by size in January 2023, new cases since 2018 and 2022; median date of first VL, baseline CD4+, baseline VL, age at first antiretroviral therapy (ART), age 2023; % key population; % regional health authorities (**Table S8**).

**Table S8. Characteristics of active phylogenetic clusters (minimum 1 new case since January 1, 2018) and were size 10 or larger**. Percentage key population and health authority are relative to reported.

| | | New cases | | | Median | | | | | Key population | | | | | Health Authority | | | | | |
|---|---|---|---|---|---|---|---|---|---|---|---|---|---|---|---|---|---|---|---|---|
| ID | Size | Since 2018 | Since 2018, PrEP | Since 2022 | Date first VL | CD4 base | VL base | Age first ART | Age in 2023 | % report | % GBM | % HET | % PWID | % HCV | % report | % Van. Coastal | % Van. Island | % Fraser | % North | % Interior |
| 57 | 471 | 19 | 0 | 5 | 2003-11-13 | 250 | 93771 | 39 | 58 | 82.0 | 5.7 | 45.9 | 85.8 | 78.2 | 94.5 | 36.4 | 20.0 | 20.4 | 19.1 | 4.0 |
| 49 | 459 | 11 | 0 | 1 | 2000-06-05 | 251 | 55200 | 40 | 60 | 85.6 | 4.8 | 47.3 | 90.8 | 87.1 | 94.3 | 55.4 | 7.6 | 20.1 | 9.2 | 7.6 |
| 31 | 391 | 94 | 8 | 11 | 2014-04-07 | 390 | 94537 | 35 | 44 | 73.9 | 88.9 | 11.4 | 16.6 | 12.5 | 94.6 | 60.5 | 19.5 | 18.6 | 0.3 | 1.1 |
| 95 | 158 | 27 | 6 | 5 | 2014-11-23 | 458 | 63537 | 35 | 42 | 63.9 | 92.1 | 8.9 | 17.8 | 12.3 | 98.1 | 63.9 | 3.9 | 31.6 | 0.6 | 0.0 |
| 13 | 150 | 3 | 0 | 0 | 2008-09-26 | 330 | 86600 | 39 | 54 | 66.0 | 88.9 | 7.1 | 13.1 | 10.4 | 97.3 | 76.7 | 6.2 | 15.8 | 0.7 | 0.7 |
| 137 | 90 | 26 | 0 | 1 | 2008-04-20 | 260 | 100010 | 37 | 53 | 84.4 | 0.0 | 47.4 | 88.2 | 82.8 | 90.0 | 70.4 | 4.9 | 17.3 | 6.2 | 1.2 |
| 142 | 62 | 6 | 0 | 2 | 2012-03-13 | 290 | 33447 | 43 | 55 | 71.0 | 75.0 | 27.3 | 20.5 | 26.3 | 88.7 | 32.7 | 56.4 | 7.3 | 0.0 | 3.6 |
| 201 | 62 | 2 | 0 | 0 | 1998-04-24 | 270 | 37000 | 38 | 60 | 90.3 | 8.9 | 55.4 | 89.3 | 89.7 | 98.4 | 65.6 | 3.3 | 16.4 | 3.3 | 11.5 |
| 175 | 56 | 1 | 0 | 0 | 2005-07-08 | 200 | 44897 | 39 | 56 | 91.1 | 2.0 | 21.6 | 98.0 | 100.0 | 92.9 | 23.1 | 3.8 | 11.5 | 57.7 | 3.8 |
| 29 | 44 | 1 | 0 | 0 | 2002-09-05 | 175 | 100010 | 42 | 59 | 79.5 | 0.0 | 42.9 | 91.4 | 87.5 | 88.6 | 71.8 | 0.0 | 23.1 | 2.6 | 2.6 |
| 22 | 42 | 19 | 5 | 7 | 2017-04-02 | 455 | 97000 | 39 | 44 | 78.6 | 100. | 0.0 | 12.1 | 10.8 | 90.5 | 73.7 | 2.6 | 21.1 | 0.0 | 2.6 |
| 72 | 40 | 3 | 0 | 0 | 2013-06-24 | 420 | 39761 | 37 | 49 | 50.0 | 80.0 | 45.0 | 15.0 | 10.3 | 97.5 | 64.1 | 0.0 | 23.1 | 2.6 | 10.3 |
| 234 | 39 | 6 | 1 | 0 | 2015-04-03 | 270 | 100010 | 33 | 44 | 84.6 | 78.8 | 18.2 | 15.2 | 13.2 | 100.0 | 30.8 | 12.8 | 7.7 | 0.0 | 48.7 |
| 73 | 37 | 2 | 0 | 0 | 2009-11-16 | 305 | 90225 | 40 | 52 | 64.9 | 87.5 | 12.5 | 16.7 | 21.2 | 91.9 | 44.1 | 14.7 | 35.3 | 0.0 | 5.9 |
| 177 | 37 | 1 | 0 | 0 | 2007-07-06 | 255 | 59000 | 41 | 53 | 89.2 | 15.2 | 51.5 | 90.9 | 88.9 | 97.3 | 69.4 | 2.8 | 25.0 | 0.0 | 2.8 |
| 109 | 36 | 1 | 0 | 0 | 1999-04-10 | 210 | 59800 | 44 | 63 | 88.9 | 6.3 | 46.9 | 96.9 | 96.9 | 94.4 | 58.8 | 23.5 | 14.7 | 2.9 | 0.0 |
| 219 | 36 | 4 | 0 | 0 | 2006-06-14 | 270 | 140000 | 40 | 55 | 75.0 | 70.4 | 29.6 | 22.2 | 18.2 | 94.4 | 61.8 | 0.0 | 38.2 | 0.0 | 0.0 |
| 102 | 33 | 1 | 0 | 0 | 2002-09-07 | 315 | 67850 | 40 | 57 | 69.7 | 82.6 | 26.1 | 26.1 | 21.2 | 97.0 | 62.5 | 25.0 | 12.5 | 0.0 | 0.0 |
| 206 | 33 | 2 | 0 | 0 | 2002-02-09 | 190 | 98405 | 36 | 56 | 84.8 | 14.3 | 60.7 | 85.7 | 79.3 | 87.9 | 69.0 | 3.4 | 24.1 | 0.0 | 3.4 |
| 217 | 33 | 4 | 0 | 0 | 2011-08-25 | 245 | 83707 | 37 | 48 | 69.7 | 0.0 | 56.5 | 60.9 | 42.9 | 90.9 | 10.0 | 0.0 | 6.7 | 80.0 | 3.3 |
| 122 | 30 | 2 | 0 | 0 | 2006-06-17 | 330 | 37847 | 37 | 53 | 90.0 | 3.7 | 29.6 | 92.6 | 89.7 | 96.7 | 20.7 | 58.6 | 20.7 | 0.0 | 0.0 |
| 41 | 29 | 7 | 2 | 0 | 2013-12-02 | 510 | 50370 | 34 | 41 | 69.0 | 80.0 | 10.0 | 25.0 | 16.0 | 93.1 | 40.7 | 33.3 | 14.8 | 3.7 | 7.4 |
| 93 | 28 | 1 | 0 | 0 | 2012-08-15 | 360 | 109908 | 29 | 42 | 60.7 | 94.1 | 0.0 | 17.6 | 8.0 | 96.4 | 70.4 | 0.0 | 18.5 | 7.4 | 3.7 |
| 26 | 26 | 5 | 2 | 0 | 2003-07-29 | 260 | 100010 | 31 | 46 | 76.9 | 85.0 | 5.0 | 20.0 | 11.5 | 100.0 | 96.2 | 0.0 | 3.8 | 0.0 | 0.0 |
| 91 | 25 | 1 | 0 | 0 | 2011-05-12 | 323 | 97567 | 37 | 48 | 52.0 | 69.2 | 7.7 | 38.5 | 20.0 | 96.0 | 54.2 | 4.2 | 37.5 | 0.0 | 4.2 |
| 209 | 25 | 5 | 0 | 2 | 2016-09-15 | 275 | 46744 | 38 | 44 | 92.0 | 4.3 | 39.1 | 78.3 | 84.0 | 92.0 | 65.2 | 4.3 | 17.4 | 4.3 | 8.7 |
| 218 | 24 | 1 | 0 | 0 | 2011-10-21 | 295 | 41503 | 35 | 45 | 70.8 | 0.0 | 35.3 | 88.2 | 80.0 | 75.0 | 5.6 | 5.6 | 5.6 | 83.3 | 0.0 |
| 132 | 22 | 1 | 0 | 0 | 2000-12-02 | 185 | 53600 | 45 | 61 | 86.4 | 0.0 | 36.8 | 100.0 | 95.5 | 100.0 | 81.8 | 0.0 | 18.2 | 0.0 | 0.0 |
| 14 | 19 | 1 | 0 | 0 | 2007-05-28 | 300 | 42349 | 43 | 56 | 73.7 | 85.7 | 0.0 | 35.7 | 31.6 | 94.7 | 83.3 | 0.0 | 16.7 | 0.0 | 0.0 |
| 168 | 18 | 17 | 5 | 17 | 2022-10-08 | 535 | 61200 | 44 | 45 | 72.2 | 100.0 | 0.0 | 0.0 | 0.0 | 83.3 | 80.0 | 0.0 | 20.0 | 0.0 | 0.0 |
| 35 | 17 | 2 | 0 | 0 | 2008-01-15 | 315 | 40300 | 37 | 52 | 82.4 | 92.9 | 21.4 | 14.3 | 12.5 | 100.0 | 58.8 | 5.9 | 29.4 | 5.9 | 0.0 |
| 40 | 17 | 3 | 0 | 0 | 2011-08-03 | 390 | 84705 | 41 | 58 | 88.2 | 86.7 | 40.0 | 13.3 | 18.8 | 82.4 | 57.1 | 14.3 | 28.6 | 0.0 | 0.0 |
| 134 | 15 | 7 | 0 | 0 | 2017-07-17 | 480 | 15250 | 37 | 50 | 80.0 | 0.0 | 41.7 | 91.7 | 91.7 | 80.0 | 66.7 | 0.0 | 33.3 | 0.0 | 0.0 |
| 146 | 15 | 1 | 0 | 1 | 2013-01-23 | 280 | 70789 | 38 | 46 | 53.3 | 75.0 | 25.0 | 12.5 | 8.3 | 80.0 | 41.7 | 16.7 | 33.3 | 8.3 | 0.0 |
| 194 | 15 | 5 | 0 | 2 | 2013-03-13 | 435 | 47245 | 36 | 44 | 86.7 | 15.4 | 61.5 | 92.3 | 85.7 | 93.3 | 78.6 | 0.0 | 21.4 | 0.0 | 0.0 |
| 221 | 15 | 1 | 0 | 0 | 2007-08-02 | 250 | 37072 | 35 | 49 | 66.7 | 100.0 | 0.0 | 10.0 | 21.4 | 93.3 | 78.6 | 14.3 | 7.1 | 0.0 | 0.0 |
| 52 | 14 | 4 | 0 | 0 | 2008-10-30 | 440 | 2662 | 36 | 44 | 71.4 | 70.0 | 60.0 | 30.0 | 7.1 | 100.0 | 57.1 | 0.0 | 35.7 | 7.1 | 0.0 |
| 114 | 14 | 14 | 2 | 6 | 2021-10-09 | 400 | 136000 | 23 | 25 | 100.0 | 92.9 | 7.1 | 7.1 | 0.0 | 92.9 | 30.8 | 15.4 | 30.8 | 0.0 | 23.1 |
| 117 | 14 | 2 | 0 | 0 | 2009-11-30 | 467 | 37289 | 43 | 59 | 71.4 | 90.0 | 40.0 | 30.0 | 21.4 | 92.9 | 30.8 | 0.0 | 61.5 | 0.0 | 7.7 |
| 193 | 13 | 3 | 0 | 0 | 2004-07-05 | 300 | 78413 | 41 | 65 | 84.6 | 0.0 | 36.4 | 100.0 | 100.0 | 76.9 | 80.0 | 0.0 | 20.0 | 0.0 | 0.0 |
| 196 | 13 | 3 | 0 | 0 | 2015-02-02 | 260 | 209844 | 34 | 47 | 92.3 | 0.0 | 83.3 | 58.3 | 38.5 | 100.0 | 53.8 | 7.7 | 38.5 | 0.0 | 0.0 |
| 202 | 13 | 9 | 0 | 1 | 2019-06-12 | 460 | 57700 | 38 | 42 | 84.6 | 0.0 | 45.5 | 100.0 | 100.0 | 92.3 | 58.3 | 0.0 | 41.7 | 0.0 | 0.0 |
| 227 | 13 | 2 | 0 | 0 | 2010-08-23 | 230 | 66535 | 28 | 39 | 69.2 | 100.0 | 11.1 | 11.1 | 8.3 | 92.3 | 41.7 | 0.0 | 58.3 | 0.0 | 0.0 |
| 245 | 13 | 1 | 0 | 0 | 2016-04-09 | 494 | 160895 | 27 | 34 | 84.6 | 81.8 | 18.2 | 9.1 | 0.0 | 100.0 | 23.1 | 0.0 | 69.2 | 0.0 | 7.7 |
| 179 | 12 | 3 | 0 | 2 | 2009-12-07 | 370 | 15945 | 36 | 45 | 75.0 | 100.0 | 11.1 | 0.0 | 0.0 | 100.0 | 8.3 | 66.7 | 16.7 | 0.0 | 8.3 |
| 197 | 12 | 5 | 1 | 2 | 2017-11-22 | 477 | 41078 | 33 | 42 | 91.7 | 81.8 | 18.2 | 0.0 | 10.0 | 91.7 | 45.5 | 0.0 | 18.2 | 0.0 | 36.4 |
| 207 | 11 | 6 | 0 | 0 | 2019-11-20 | 560 | 240000 | 36 | 42 | 90.9 | 10.0 | 30.0 | 80.0 | 81.8 | 81.8 | 22.2 | 0.0 | 55.6 | 11.1 | 11.1 |
| 33 | 10 | 1 | 0 | 0 | 2010-05-22 | 500 | 49606 | 37 | 55 | 20.0 | 100.0 | 0.0 | 0.0 | 0.0 | 80.0 | 87.5 | 0.0 | 12.5 | 0.0 | 0.0 |
| 173 | 10 | 1 | 0 | 0 | 2011-05-21 | 345 | 107236 | 41 | 51 | 80.0 | 100.0 | 0.0 | 12.5 | 22.2 | 90.0 | 77.8 | 0.0 | 11.1 | 11.1 | 0.0 |
| 182 | 10 | 3 | 0 | 0 | 2008-02-28 | 150 | 120023 | 48 | 61 | 70.0 | 28.6 | 85.7 | 28.6 | 50.0 | 80.0 | 12.5 | 0.0 | 37.5 | 50.0 | 0.0 |
| 203 | 10 | 1 | 0 | 0 | 2003-01-21 | 240 | 83005 | 40 | 55 | 90.0 | 11.1 | 44.4 | 100.0 | 88.9 | 90.0 | 66.7 | 11.1 | 11.1 | 0.0 | 11.1 |
| 205 | 10 | 9 | 0 | 2 | 2020-08-22 | 660 | 171000 | 31 | 33 | 60.0 | 0.0 | 33.3 | 100.0 | 100.0 | 80.0 | 87.5 | 0.0 | 12.5 | 0.0 | 0.0 |
| 208 | 10 | 1 | 0 | 0 | 2010-11-16 | 360 | 23149 | 40 | 51 | 80.0 | 0.0 | 50.0 | 62.5 | 60.0 | 100.0 | 50.0 | 0.0 | 50.0 | 0.0 | 0.0 |

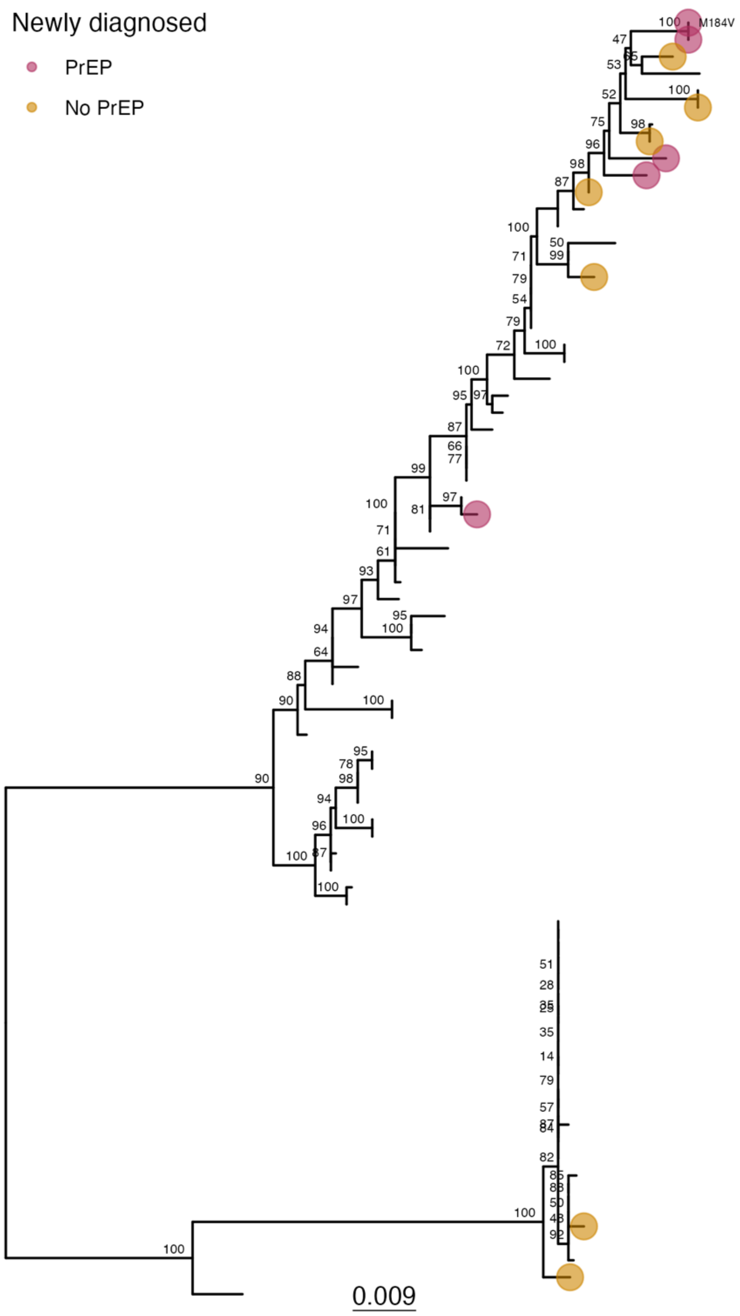


**Fig. S8. Maximum likelihood phylogenetic tree of HIV cluster 22**. Tree was inferred with partial pol alignment stripped of drug resistance mutation codons in IQTREE with a GTR model, and rooted using root-to-tip regression. Nodes annotated with ultrafast bootstrap support values. Tips colored for viruses from newly diagnosed (2018-2022) PrEP users and non-PrEP users, and annotated with NRTI treatment selected mutations including M184IV. Multiple sequences shown per participant.

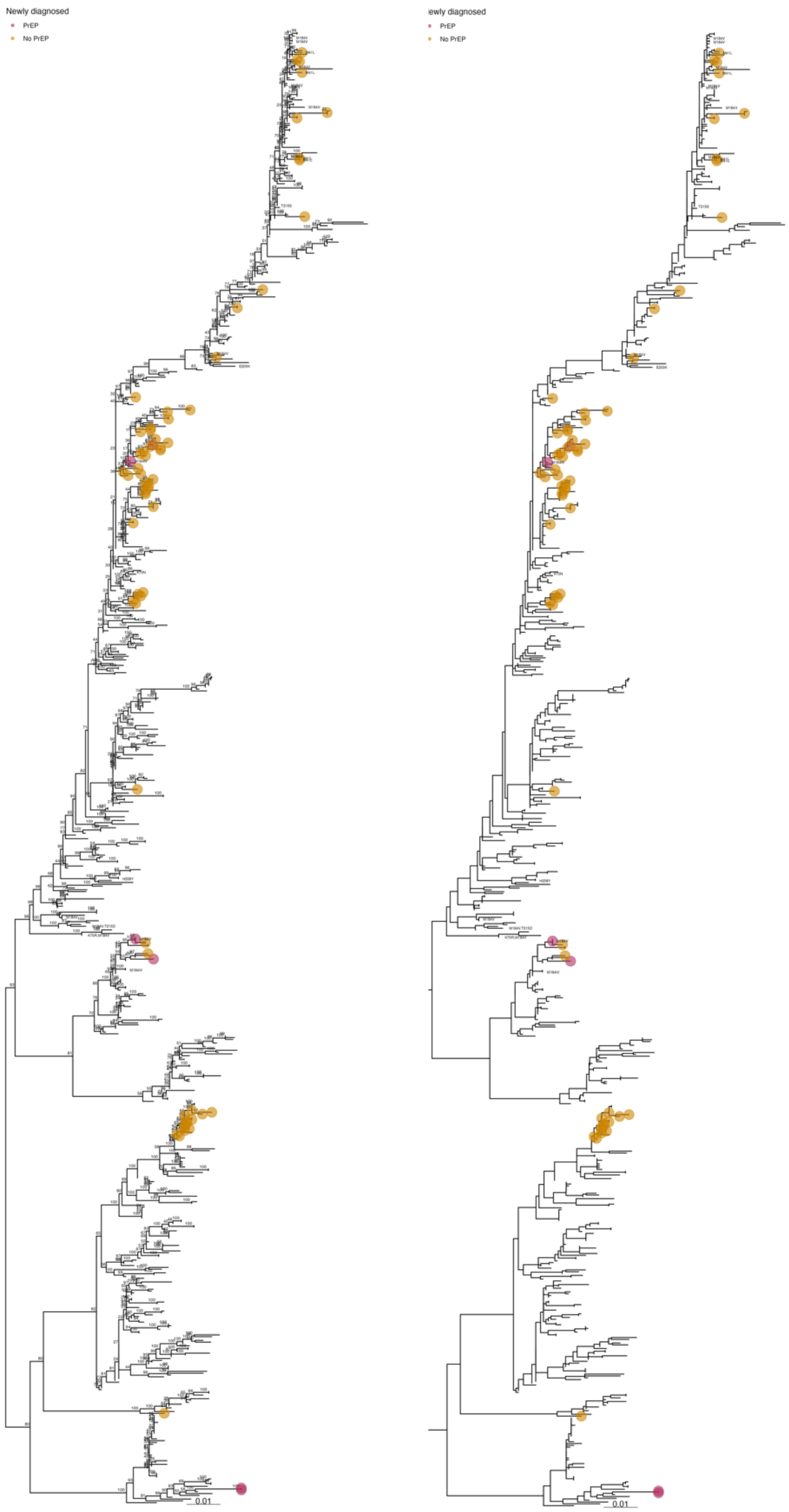

292 **Fig. S9. Maximum likelihood phylogenetic tree of HIV cluster 31, with and without support values.**

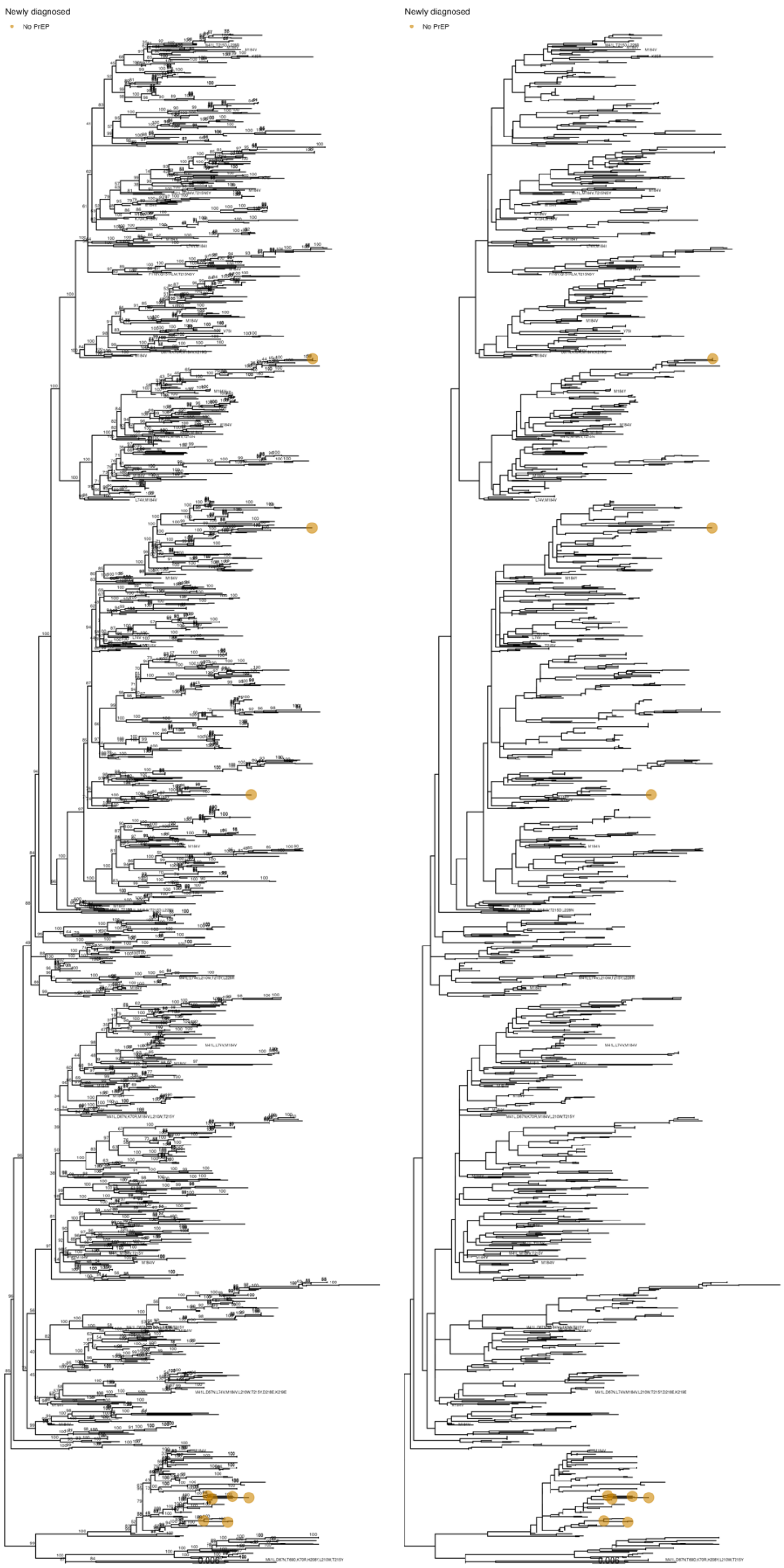


293 **Fig. S10. Maximum likelihood phylogenetic tree of HIV cluster 49, with and without support**
294 **values.**

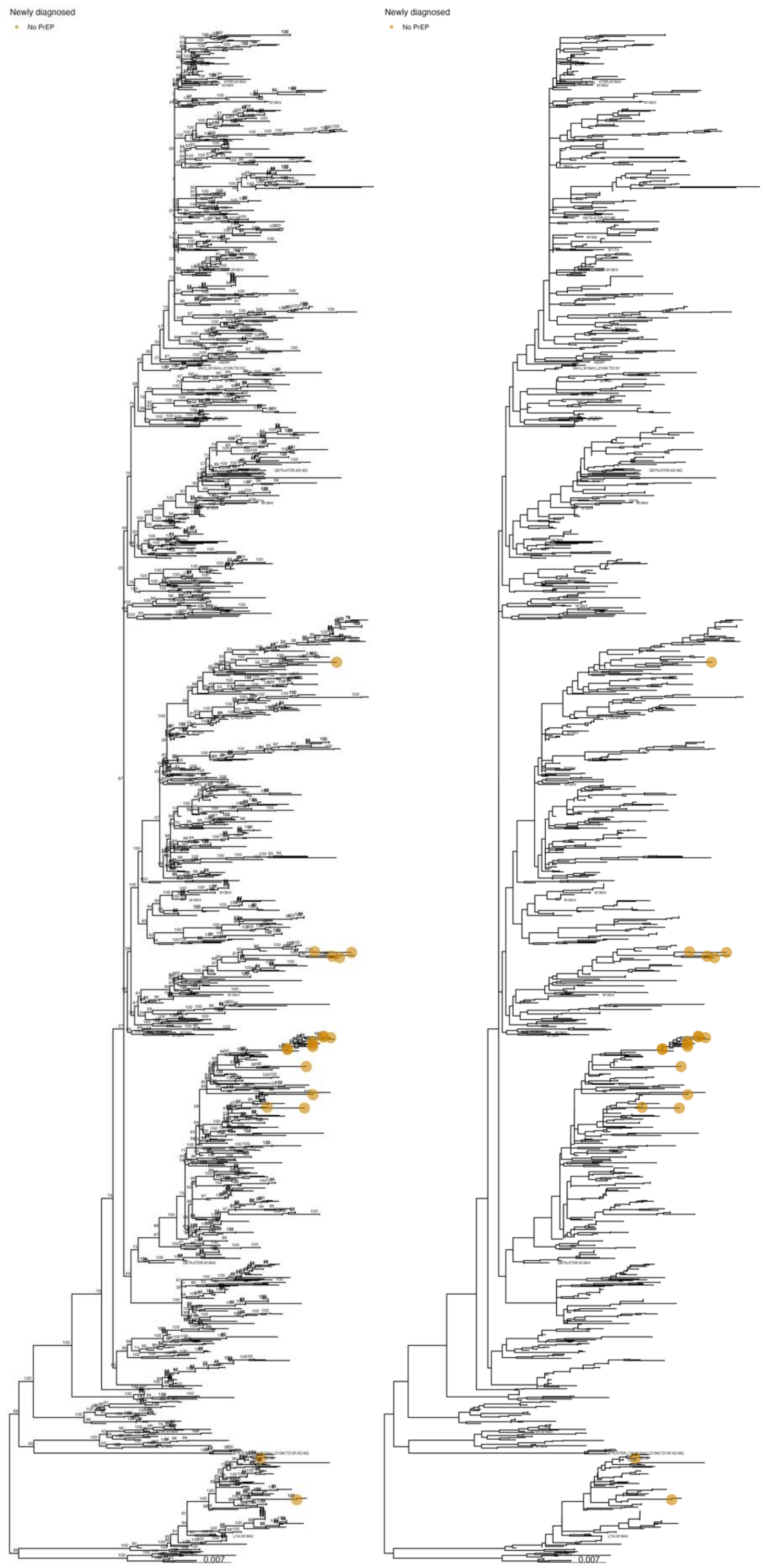


Fig. S11. Maximum likelihood phylogenetic tree of HIV cluster 57, with and without support values.

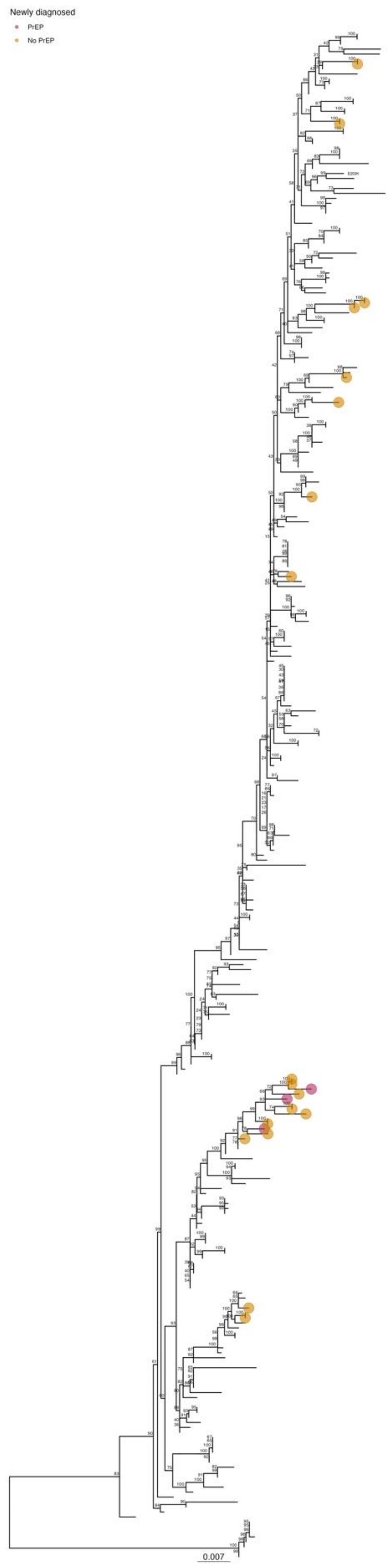


**Fig. S12. Maximum likelihood phylogenetic tree of HIV cluster 95.**

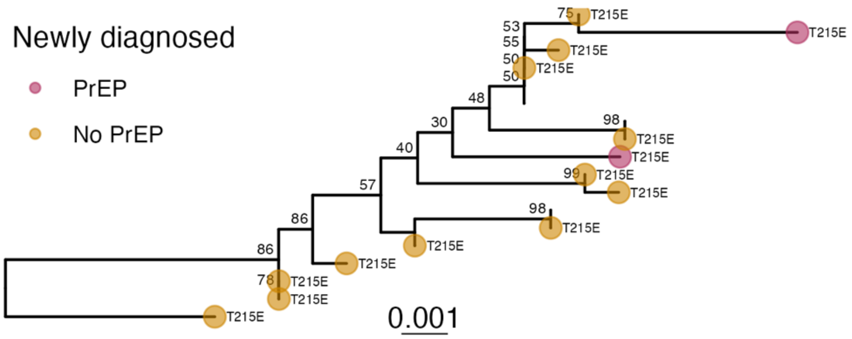


**Fig. S13. Maximum likelihood phylogenetic tree of HIV cluster 114.**

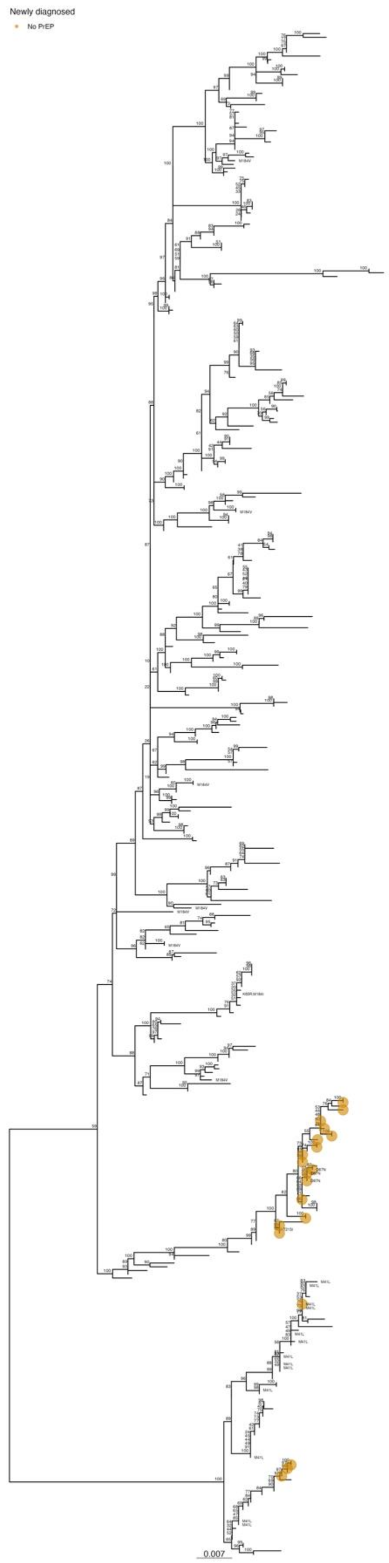


**Fig. S14. Maximum likelihood phylogenetic tree of HIV cluster 137.**

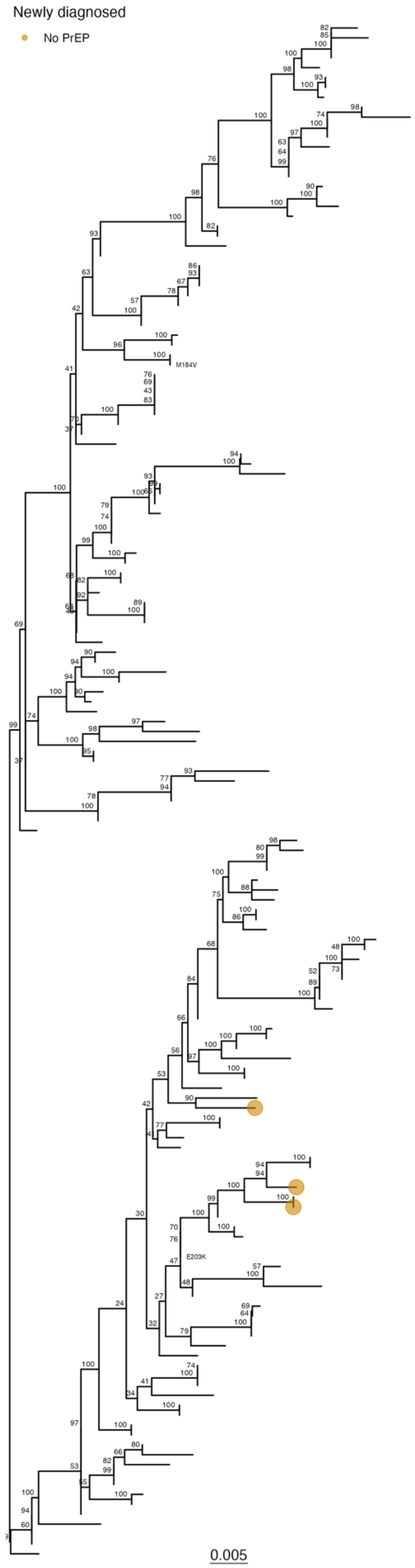


**Fig. S15. Maximum likelihood phylogenetic tree of HIV cluster 142.**

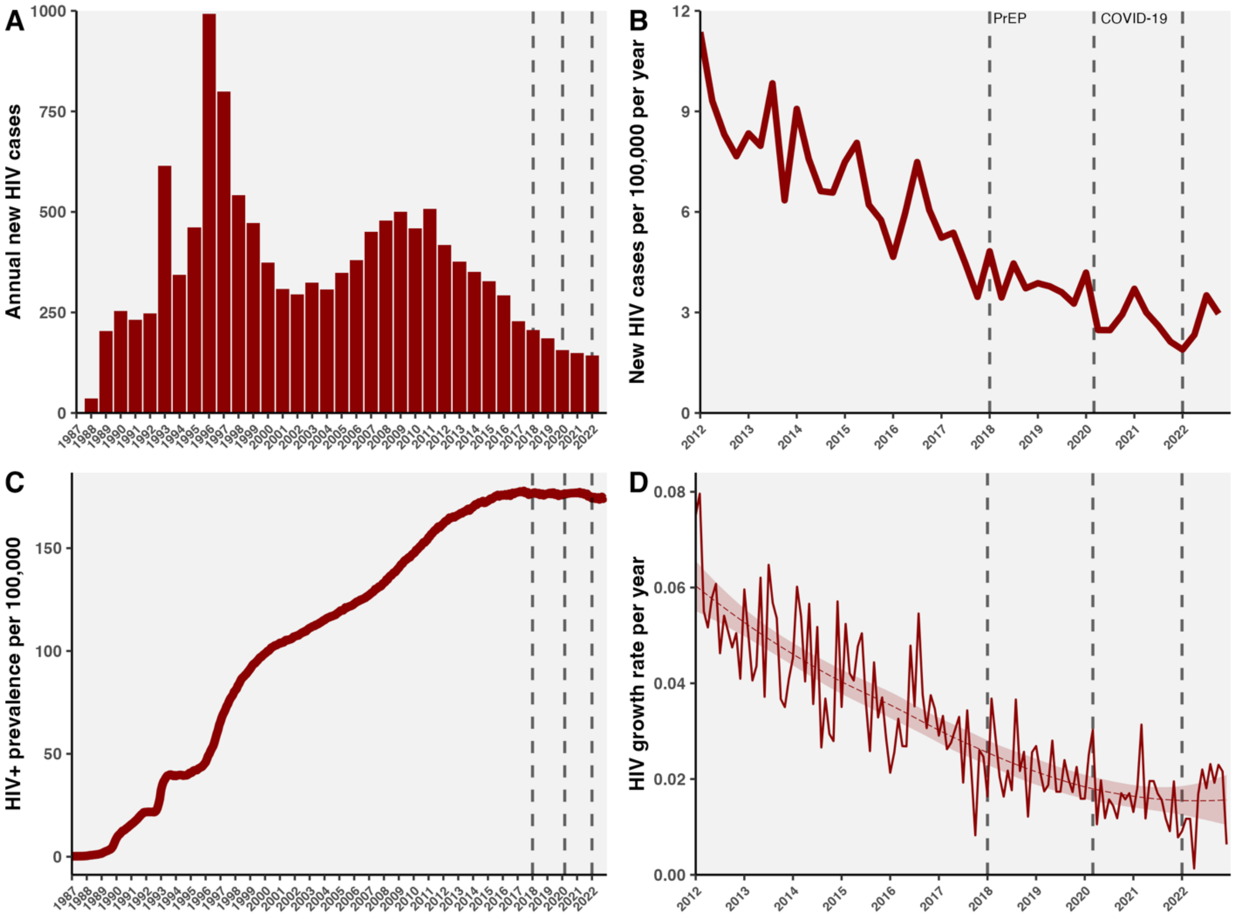


**Fig. S16. HIV incidence and prevalence in BC in the context of PrEP availability. A)** Annual new HIV cases in BC from 1987 to the end of 2022, estimated by the date of first antiretroviral treatment, excluding those with previous ART history. **B)** New HIV cases in BC per 100,000 per year calculated quarterly from 2012 to 2022, normalized to BC population size BC in 2023.[18] **C)** HIV+ prevalence per 100,000 in BC from 1987 to 2022, calculated as the cumulative sum of new cases and HIV+ migrants, subtracting the number of deaths and emigrants. **D)** HIV growth rate per year in BC from 2012 to 2022, calculated as new cases divided by the prevalent population size in each calendar month converted to per year. Dotted vertical lines denote PrEP availability in January 2018 onwards and COVID-19 pandemic interventions from March 2020 to January 2022.

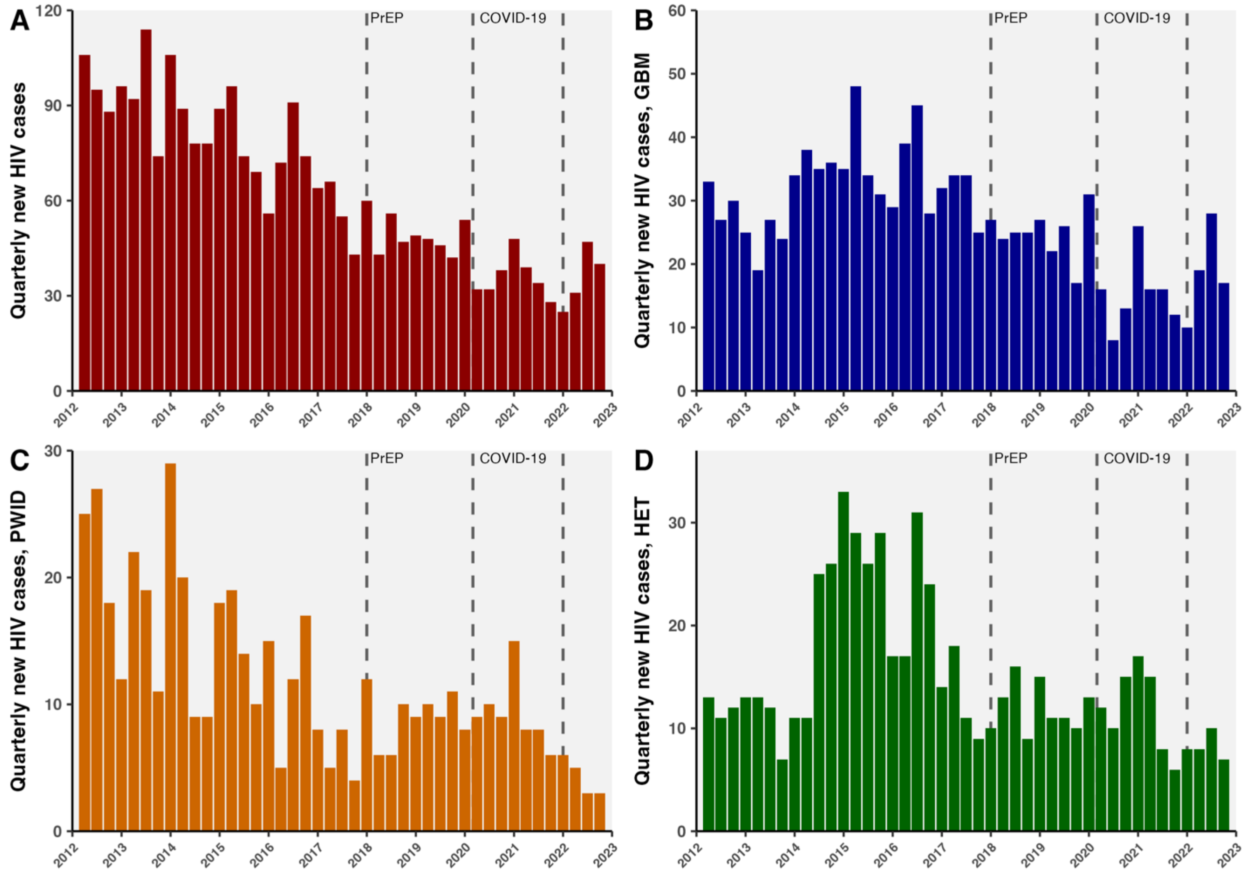


**Fig. S17. Quarterly new HIV cases from January 2012 to February 2023 by key population**. For **A)** all BC residents, **B)** gay, bisexual, and other men who have sex with men (GBM), **C)** people who inject drugs (PWID), and **D)** heterosexuals (HET). Individuals with multiple risk factors are represented in multiple panels. Dotted vertical lines denote PrEP availability in January 2018 onwards and COVID-19 pandemic interventions from March 2020 to January 2022.

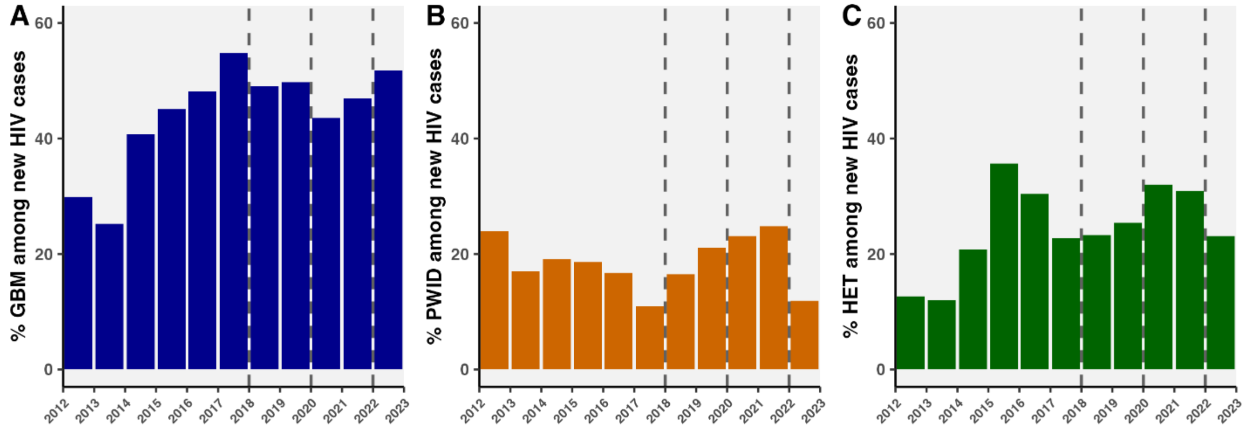


**Fig. S18. Percentage of annual new HIV cases within key populations. A)** GBM, **B)** PWID, **C)** HET. Individuals who reported multiple risk factors are represented in multiple panels and not all individuals reported risk factors. Dotted vertical lines denote PrEP availability in January 2018 onwards and COVID-19 pandemic interventions from March 2020 to January 2022.

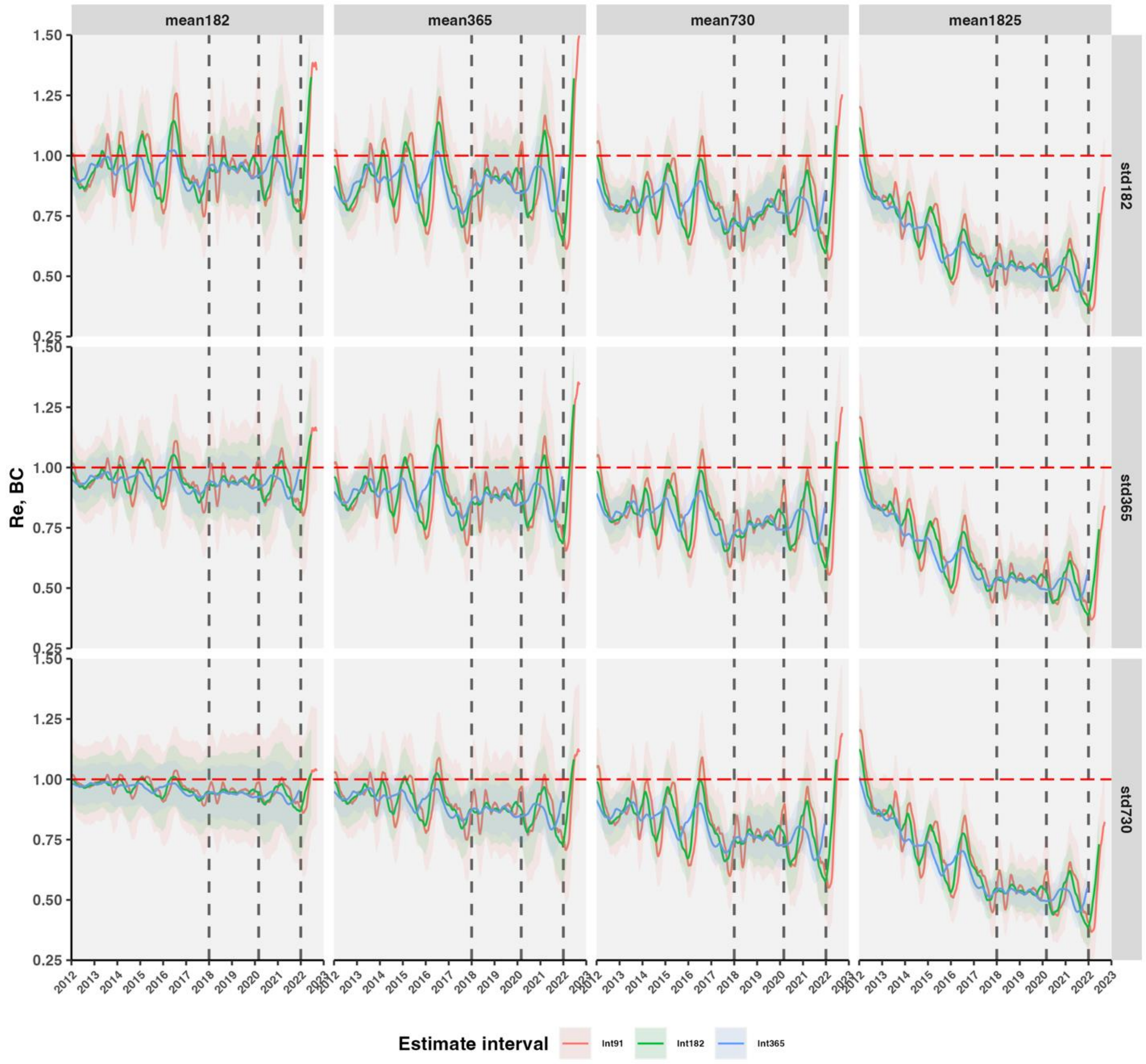


**Fig. S19. Effective reproduction number ($R_e$) of HIV in BC from 2012 to 2022 under varying serial interval assumptions.** Instantaneous $R_e$ was estimated in EpiEstim with gamma-distributed serial intervals for multiple parameter sets with means (by columns) of 0.5 year (y) (182 days(d)), 1 y (365 d), 2 y (730 d), or 5 y (1825 d); standard deviations (by rows) of 0.5 y, 1 y, or 2 y; and estimate windows (by color) of 0.25 y (91 d), 0.5 y, or 1 y. $R_e$ were smoothed with k=90 d, right-aligned (i.e. average over past 90 d).

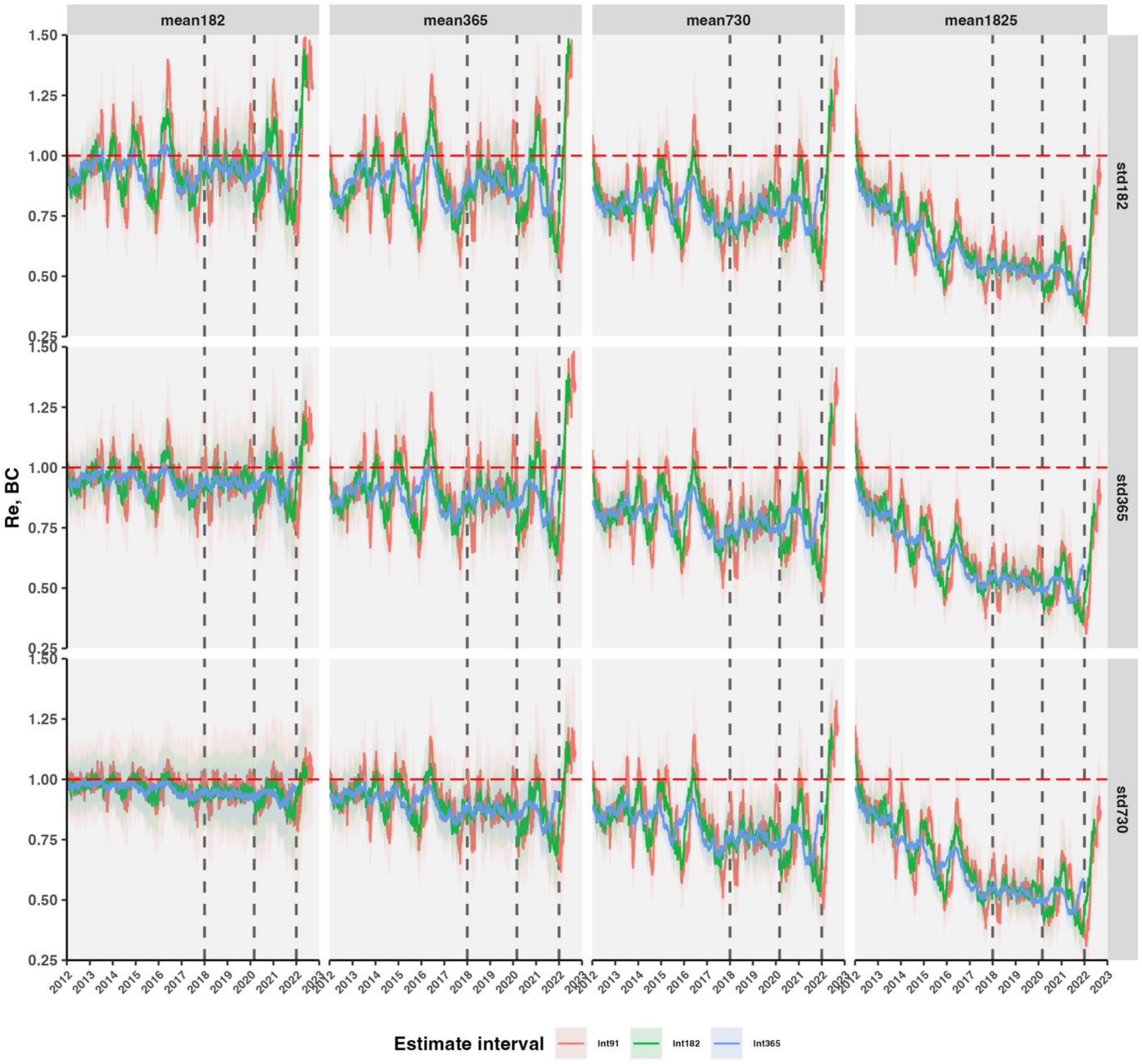


**Fig. S20. HIV effective reproductive number ($R_e$) in BC as in Fig. S19, with no smoothing.**

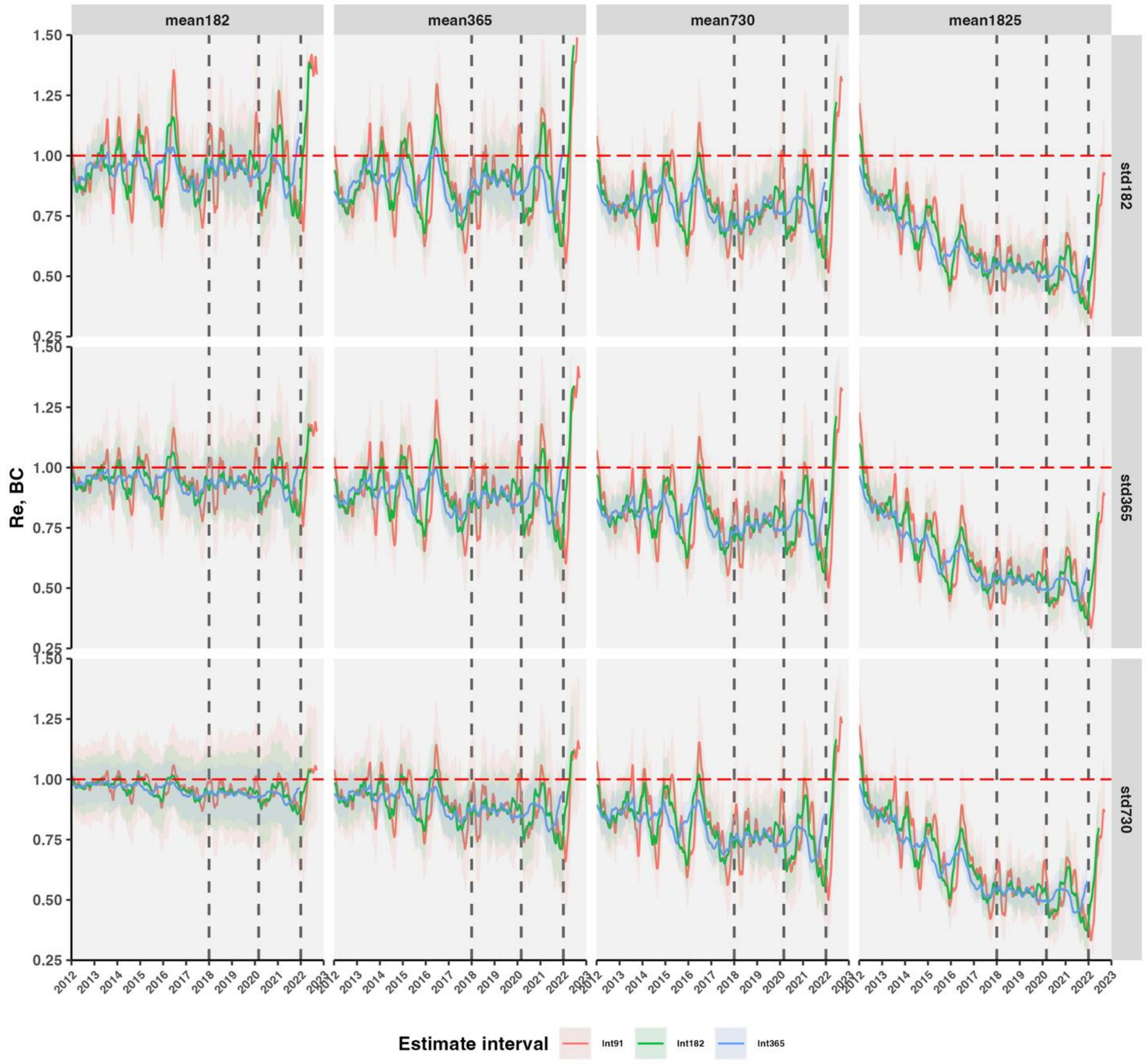


**Fig. S21. HIV effective reproductive number ($R_e$) in BC as in Fig. S19, with 30 d smoothing.**

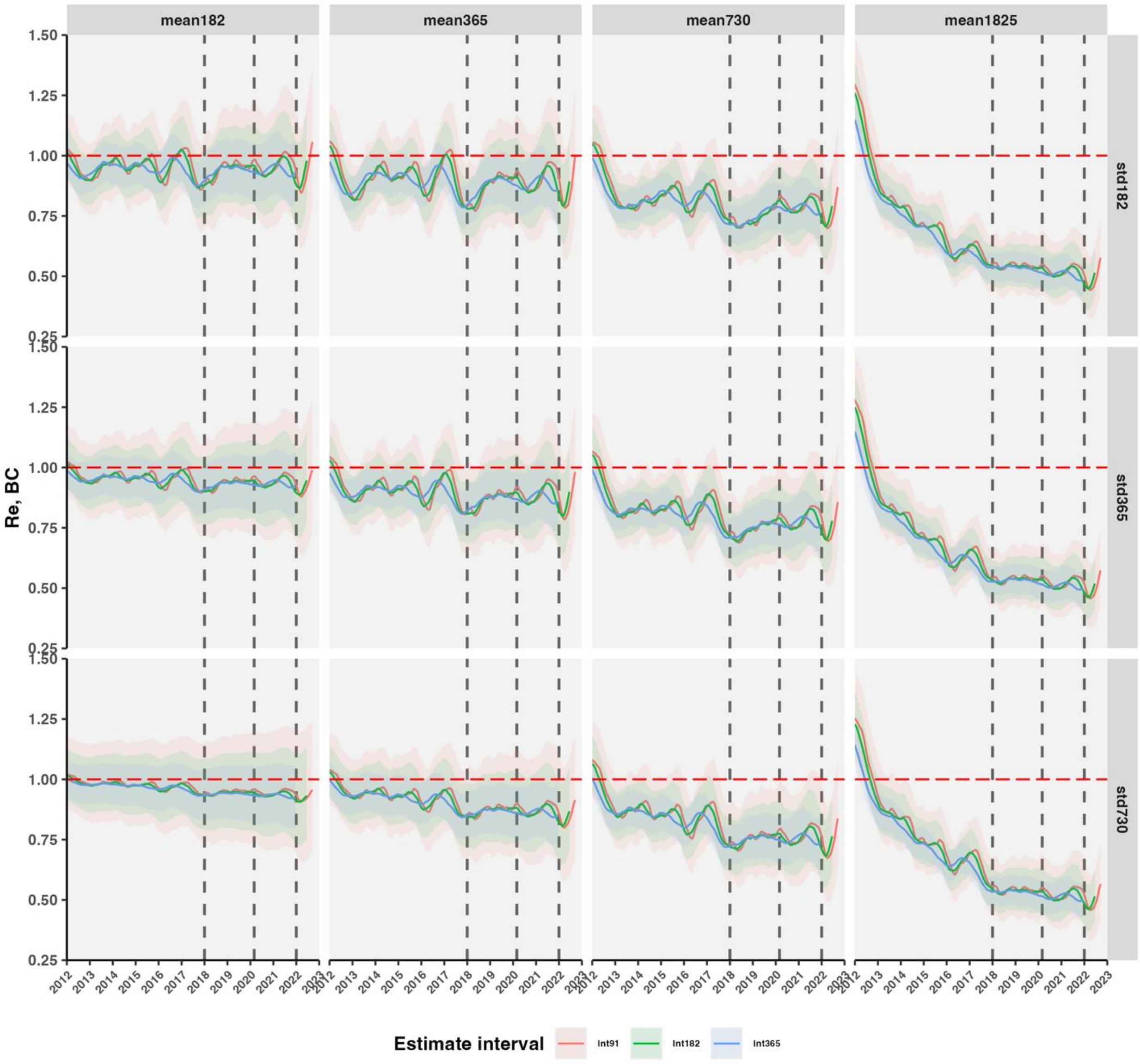


**Fig. S22. HIV effective reproductive number ($R_e$) in BC as in Fig. S19, with 365 d smoothing.**

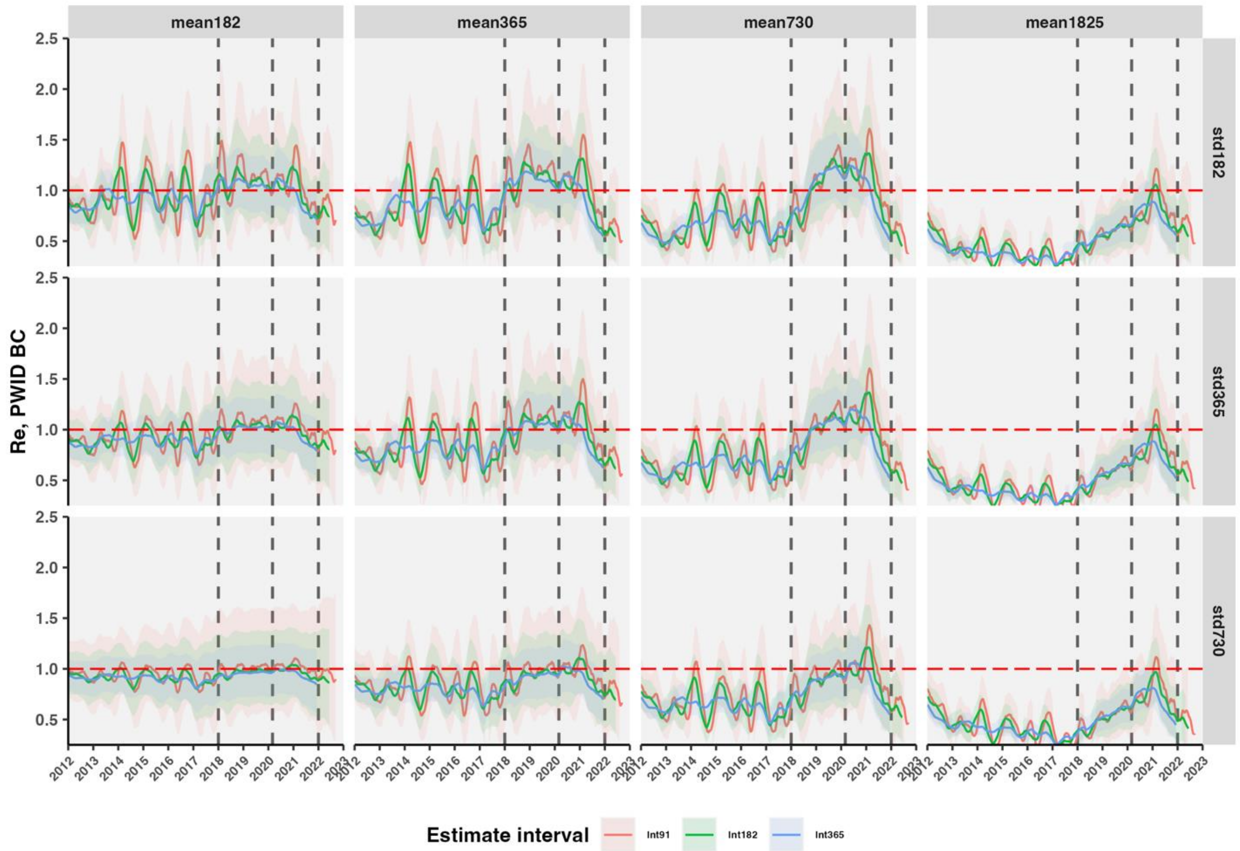


**Fig. S23. HIV effective reproductive number ($R_e$) among GBM in BC from 2012 to 2022 under multiple serial interval assumptions.** $R_e$ estimated with gamma distributed serial intervals for multiple parameters with means (by columns) of 0.5 y (182 d), 1 y (365 d), 2 y (730 d), or 5 y (1825 d); standard deviations (by rows) of 0.5 y, 1 y, or 2 y; and estimate intervals (by color) of 0.25 y (91 d), 0.5 y, or 1 y. $R_e$ were smoothed with k=90 d, right-aligned.

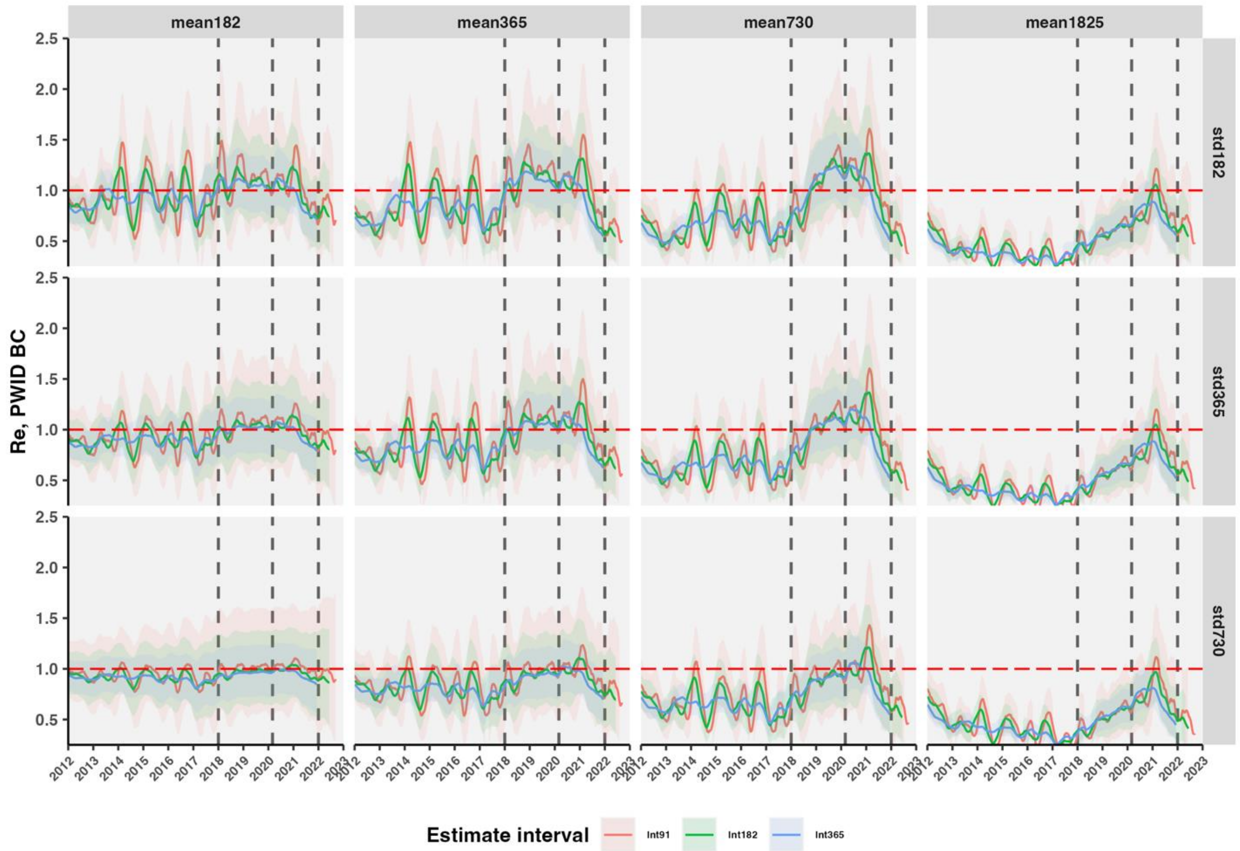


**Fig. S24. HIV effective reproductive number ($R_e$) among PWID in BC from 2012 to 2022 under multiple serial interval assumptions.** $R_e$ was estimated with gamma distributed serial intervals for multiple parameters with means (by columns) of 0.5 y (182 d), 1 y (365 d), 2 y (730 d), or 5 y (1825 d); standard deviations (by rows) of 0.5 y, 1 y, or 2 y; and estimate intervals (by color) of 0.25 y (91 d), 0.5 y, or 1 y. $R_e$ were smoothed by 90 d, right-aligned.

Longer mean serial intervals up to five years were associated with $R_e$ further from 1, and is unrealistic in recent years where most individuals have been rapidly diagnosed and untransmissible with antiretroviral treatment. Wider standard deviation of the serial interval gamma distribution brought $R_e$ closer to 1 (**Fig. S13**). Lower mean serial intervals were associated with wider confidence intervals. In our primary analysis, we report $R_e$ estimated with intermediate values: gamma-distributed serial interval with mean 1 y, standard deviation 1 y, and $R_e$ estimate interval of 0·5 y, and 90 d smoothing. Temporal trends in the $R_e$ inflection points were similar for multiple smoothing windows (**Figs. S14-S15**).

#### Phylogenetic cluster-specific effective reproduction number

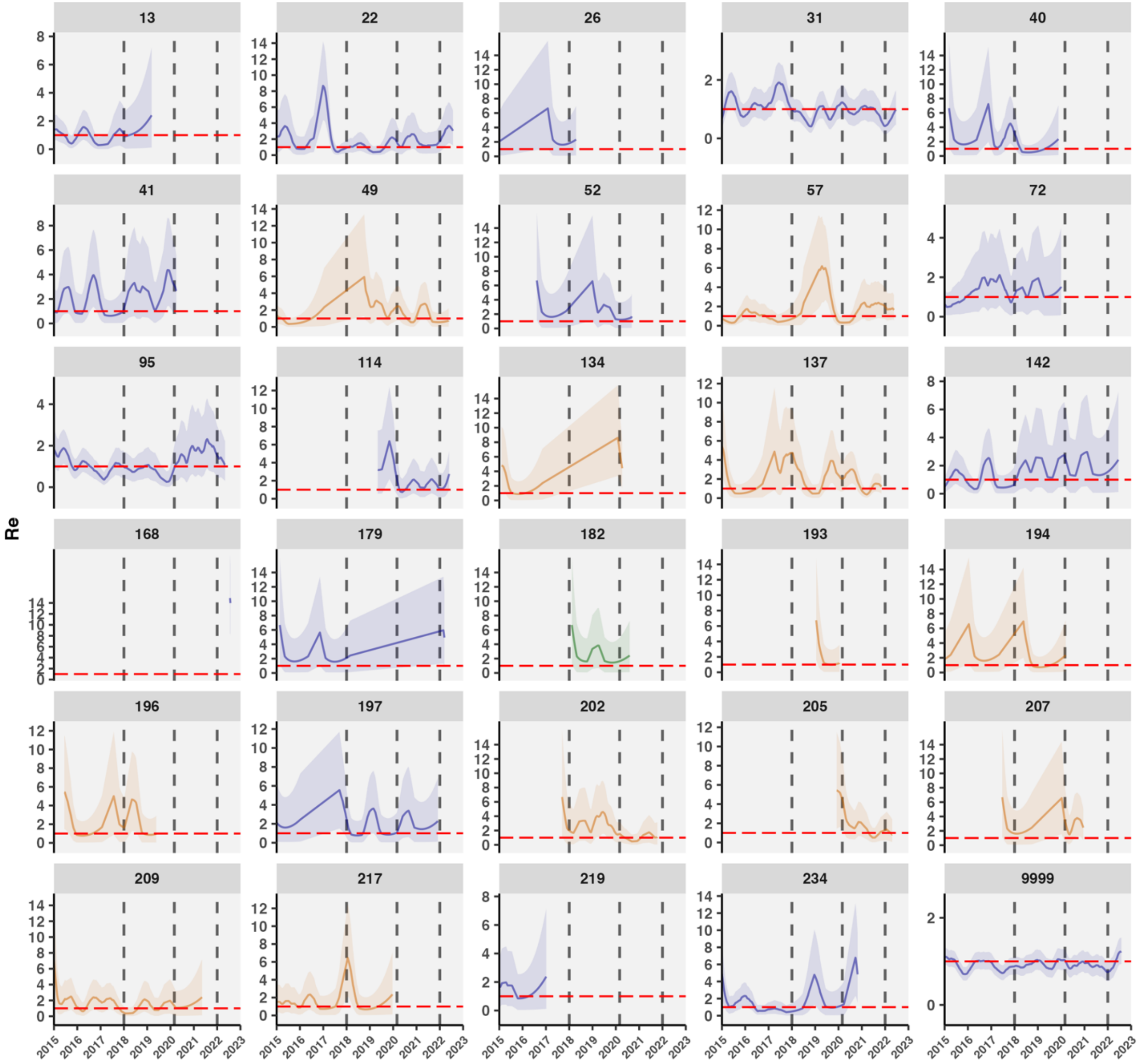


**Fig. S25. HIV effective reproductive number ($R_e$) across large (size in 2022>9) and active (new cases since 2018>2) clusters, estimated with gamma-distributed serial interval mean 1 y, standard deviation (sd) 0.5 y, estimating window 0.5 y, and 90 d smoothing.** Excludes values where the confidence interval width exceeded 10 due to sparse cases. Cluster 9999 is all non-clustered cases. Color denotes predominant population (blue: GBM, orange: PWID, and green: HET).

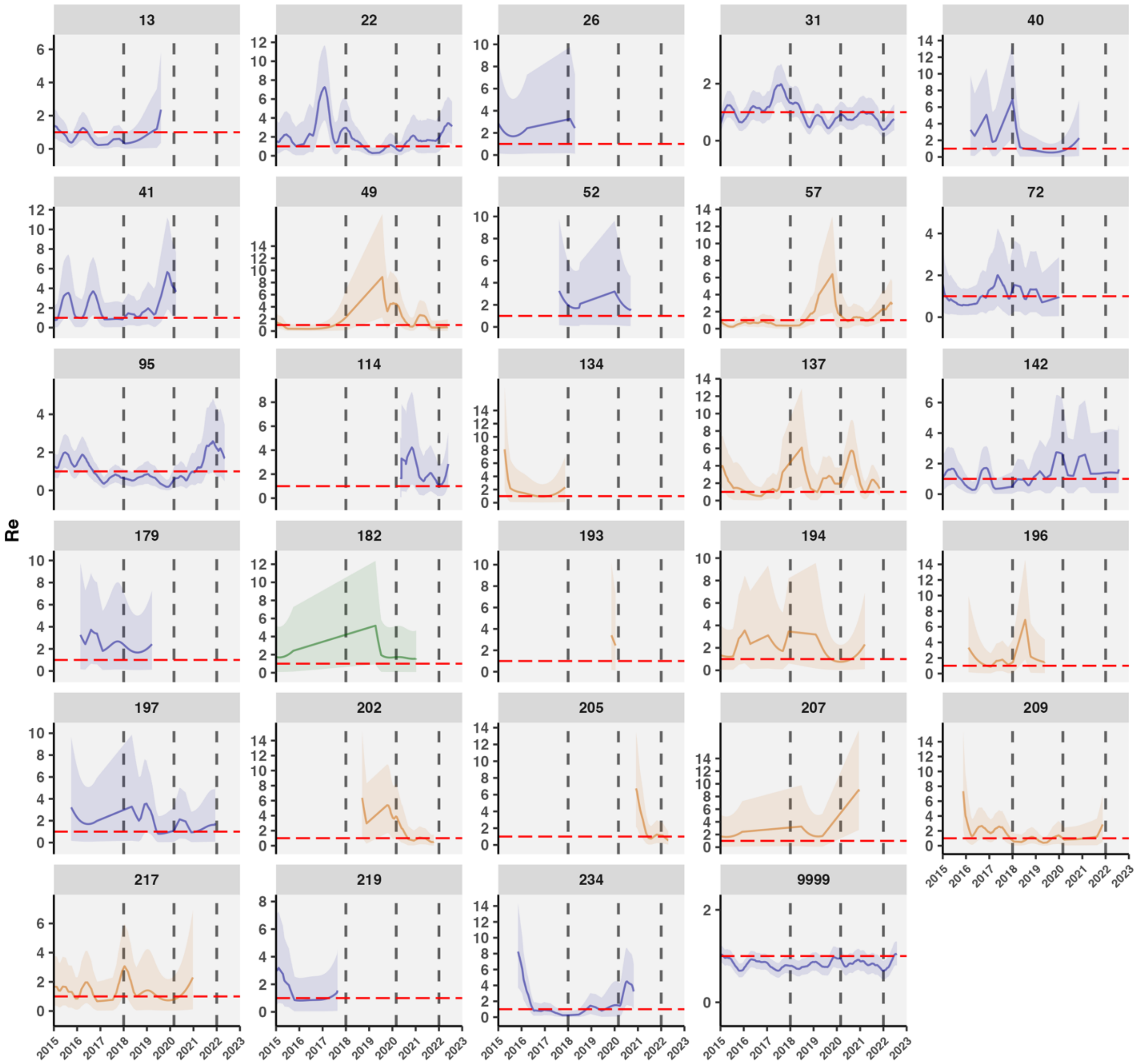


**Fig. S26. HIV effective reproductive number ($R_e$) across large and active clusters, estimated with gamma-distributed serial interval *mean 2 y*, sd 0.5 y, estimating window 0.5 y, and 90 d smoothing.**

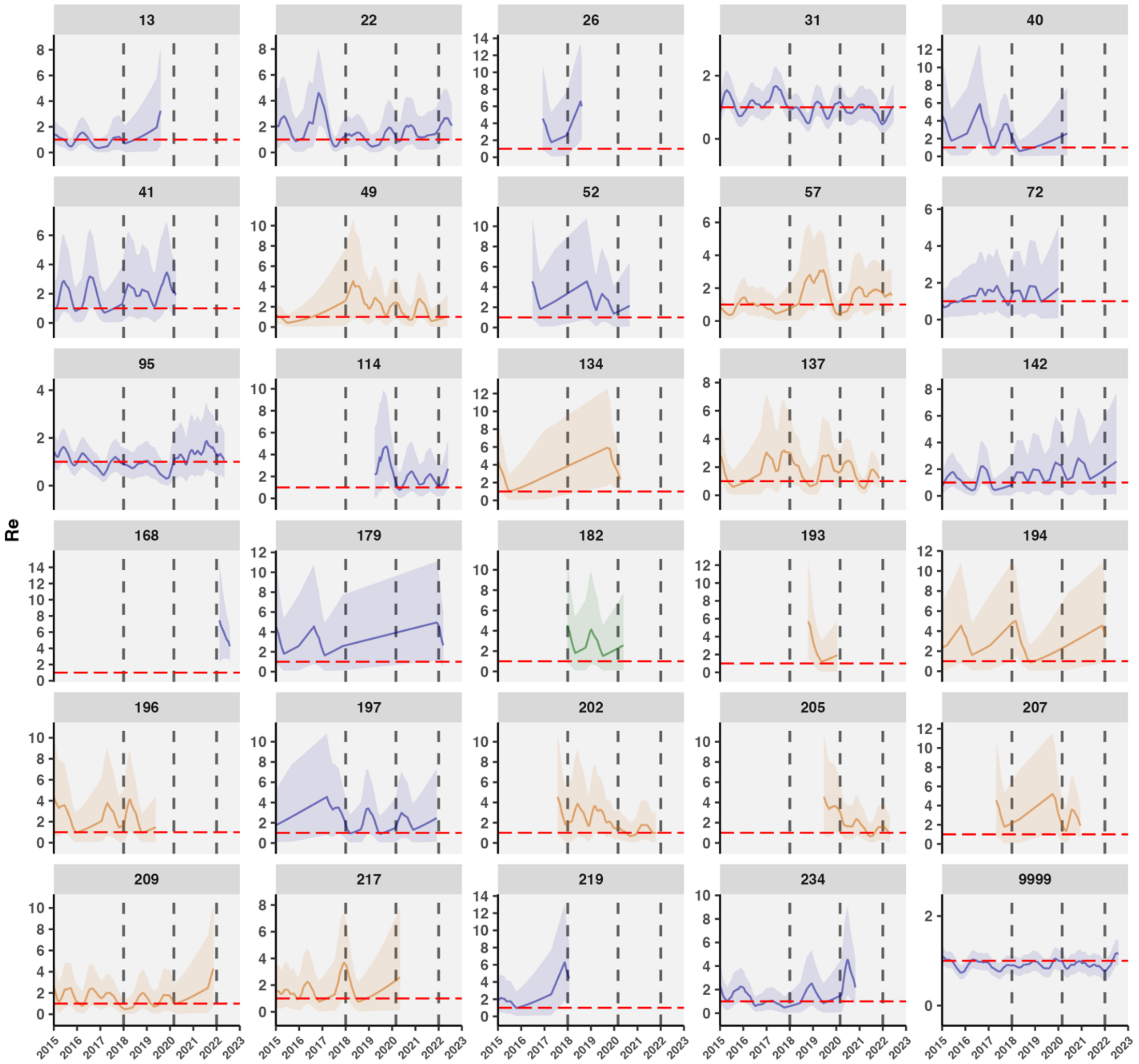


**Fig. S27. HIV effective reproductive number ($R_e$) across large and active clusters, estimated with gamma-distributed serial interval mean 1 y, *sd 1 y*, estimating window 0.5 y, and 90 d smoothing.**

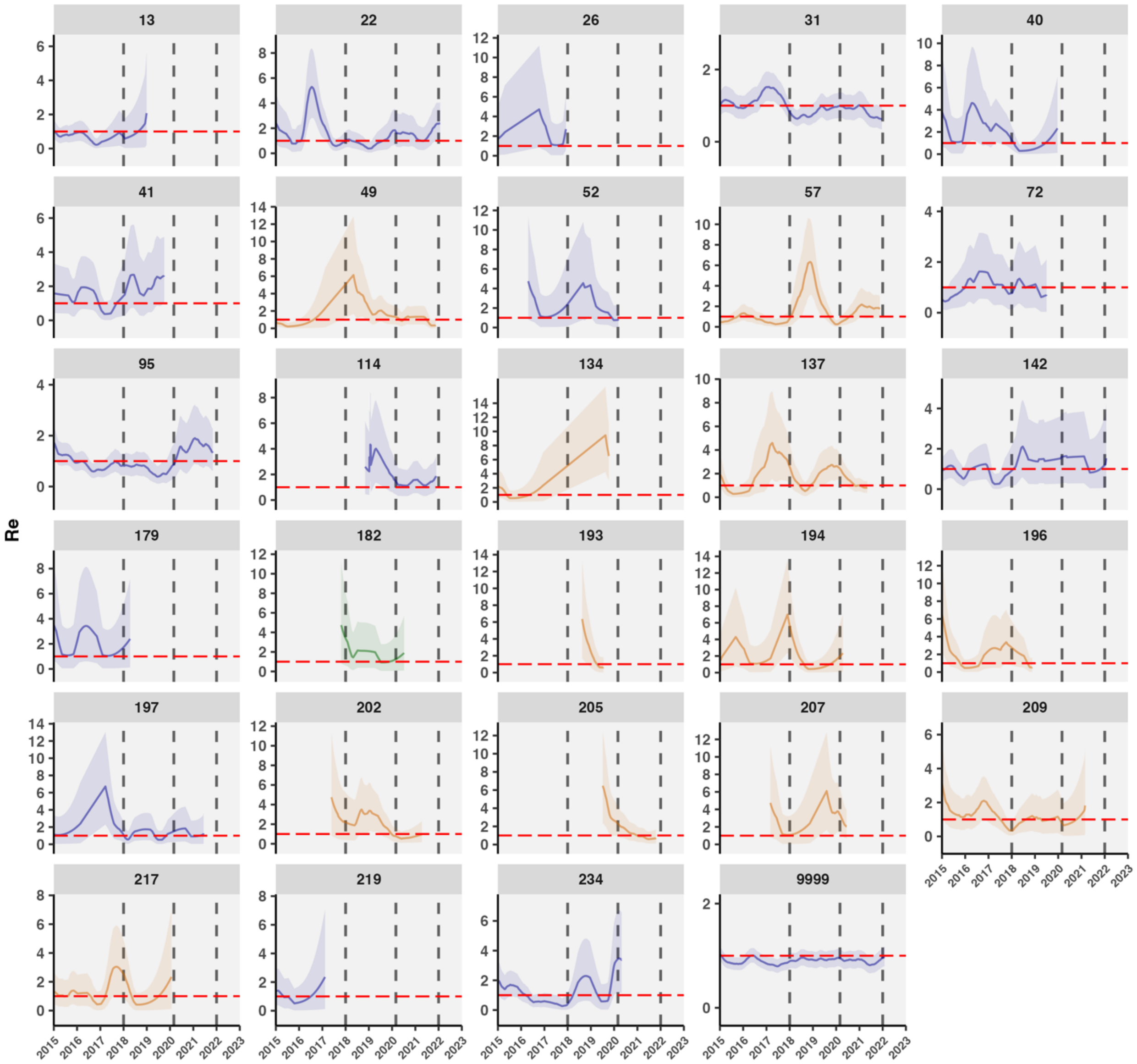


**Fig. S28. HIV effective reproductive number ($R_e$) across large and active clusters, estimated with gamma-distributed serial interval mean 1 y, sd 0.5 y, *estimating window 1 y*, and 90 d smoothing.**

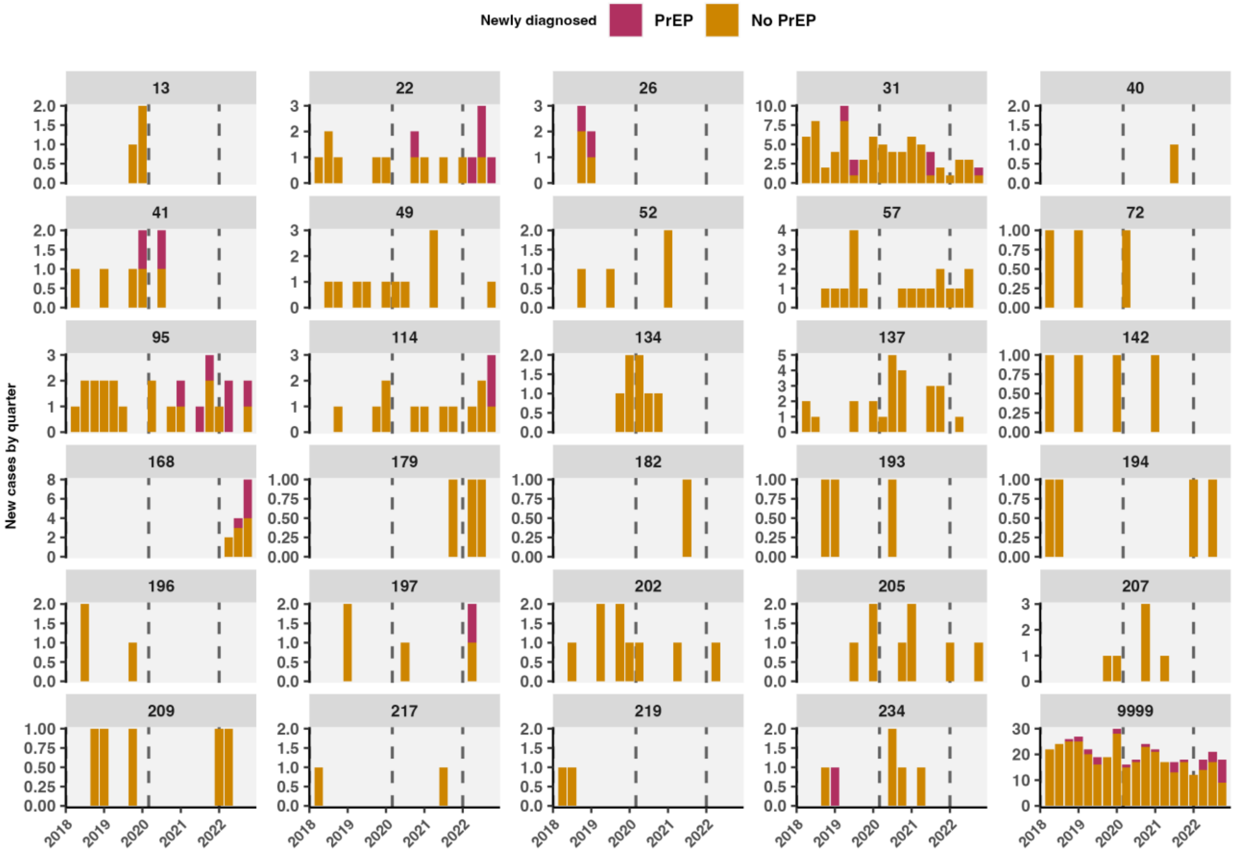


**Fig. S29. Quarterly new cases in large active clusters by PrEP use.**

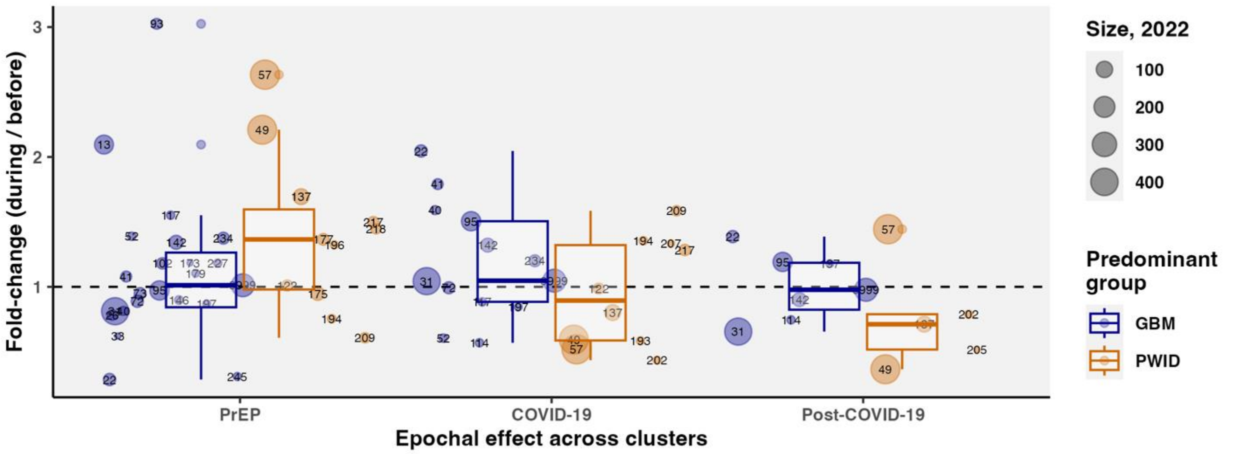


**Fig. S30. Epochal effects of PrEP and COVID-19 on cluster-level $R_e$.** Epochal effects were calculated as fold-change in the piecewise average $R_e$ in the PrEP period, compared to before PrEP (during PrEP/pre-COVID, Jan. 2018 – Feb. 2020 vs. before PrEP, Jan. 2016 – Dec. 2017), COVID-19 (during COVID, Mar. 2020 – Dec. 2021 vs. during PrEP/pre-COVID), and post-COVID-19 (Jan. – Dec. 2022 vs. during COVID). Point size represents cluster size in 2022, color is predominant population. Restricted to medium (>=10 size), active (>=1 case since 2018) clusters. Box plots show median and interquartile range of predominant groups' epochal effects.

#### Counterfactual simulations of cluster growth

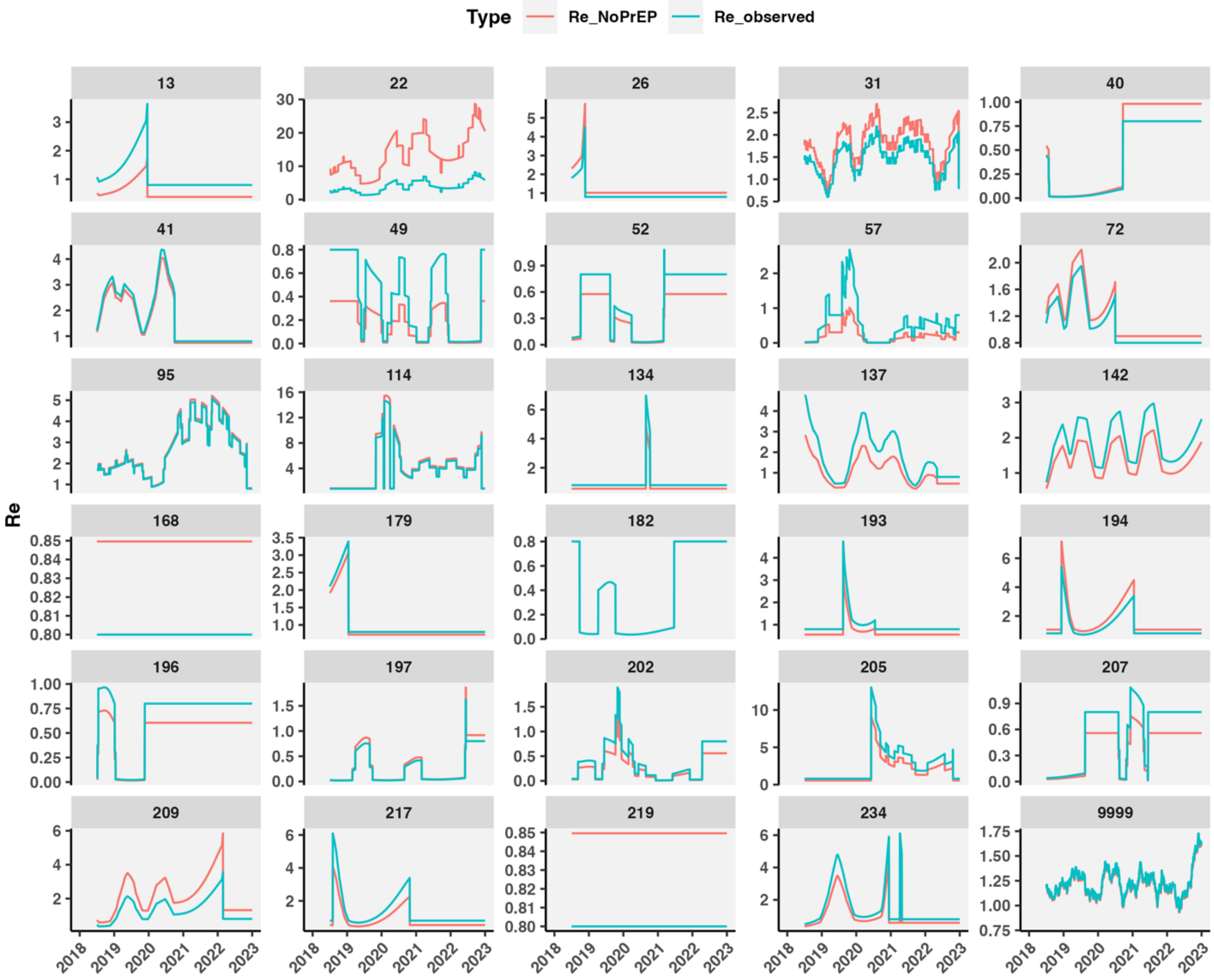


**Fig. S31. Observed and adjusted (in the absence of PrEP) cluster $R_e$.** Displaying clusters with >=3 new cases since 2018 and size in 2023 >=10.

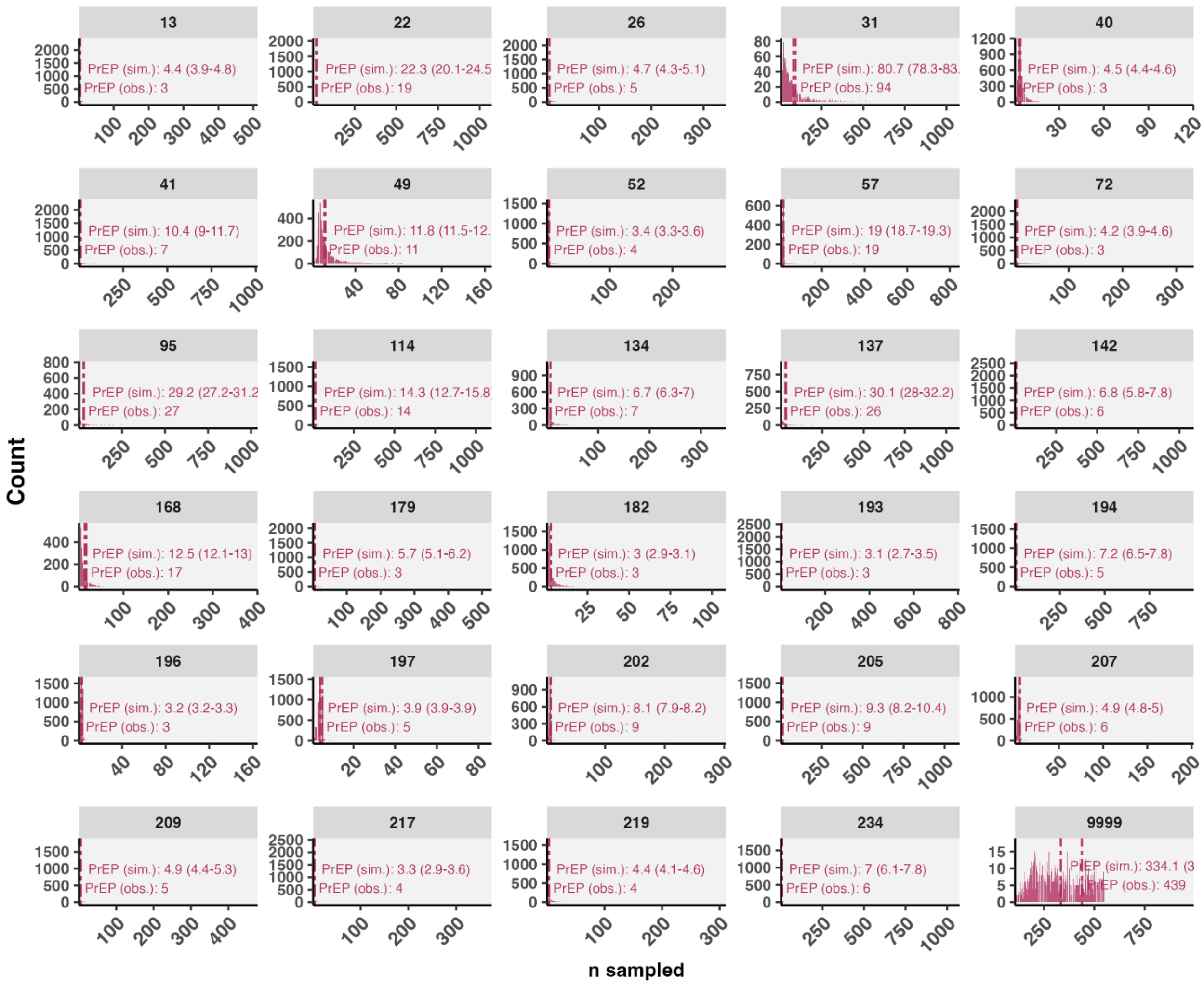


**Fig. S32. Stochastic branching processes recapitulate observed cluster growth.** Observed number of new samples ‘PrEP (obs.)’ in large, active clusters from 2018 to 2022 compared to distribution of new samples across 2000 simulations using clusters’ observed $R_e$ ‘PrEP (sim.)’.

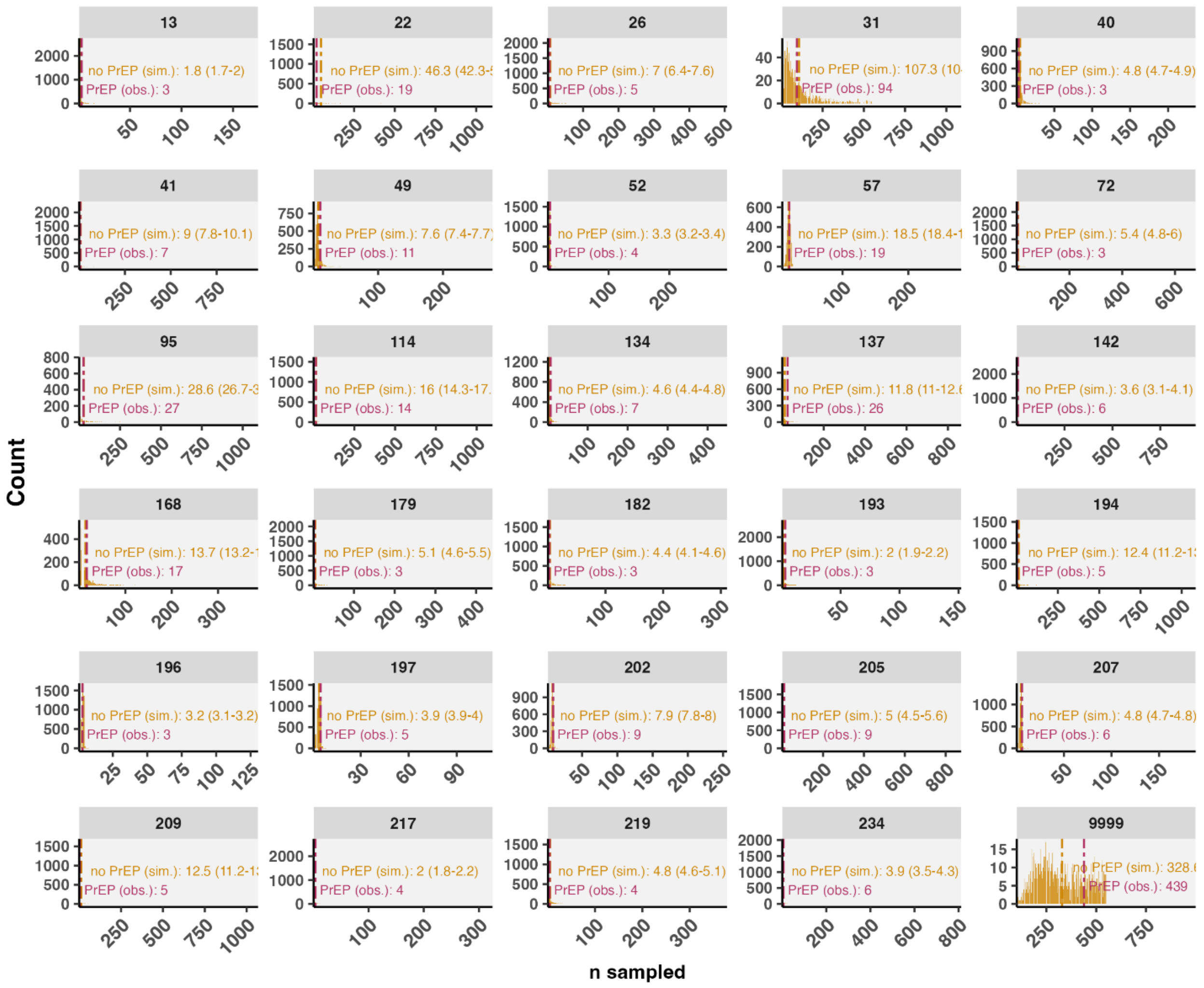


**Fig. S33. Simulations of cluster samples in the absence of PrEP.** Observed number of new samples (among new diagnoses) 'PrEP (obs.)' in large, active clusters from 2018 to 2022 compared to distribution of new samples across 4000 simulations using clusters $R_e$ adjusted by PrEP effect 'no PrEP (sim.)'.

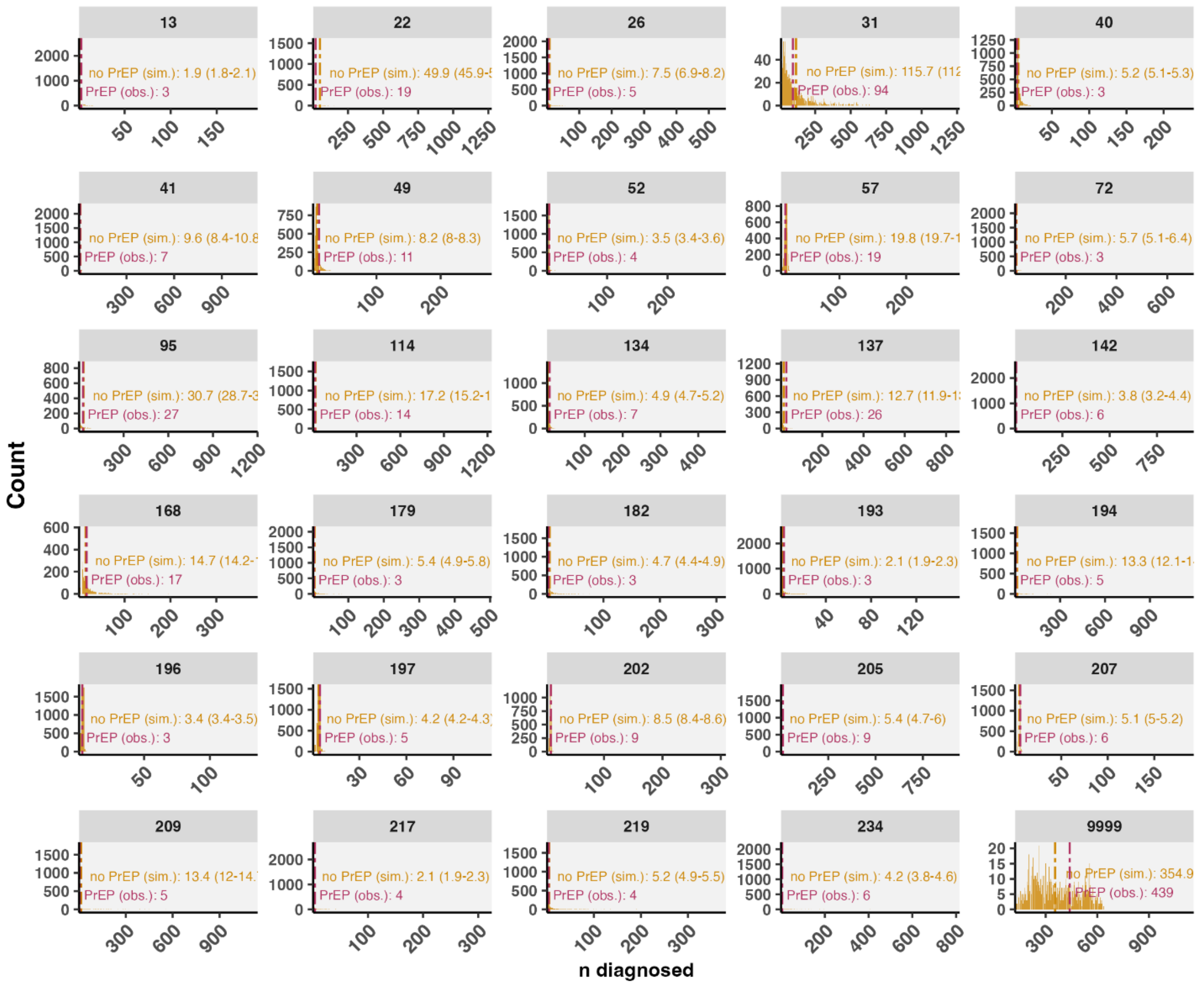


**Fig. S34. Simulations of cluster diagnoses in the absence of PrEP.** Observed number of new diagnoses 'PrEP (obs.)' in large, active clusters from 2018 to 2022 compared to distribution of new diagnoses across 4000 simulations using clusters $R_e$ adjusted by PrEP effect 'no PrEP (sim.)'.

**Table S9. Poisson model of diagnoses averted across clusters**. Counts were normalized to be positive integers (minimum averted added to all). Exponentiated coefficients reported. Significant adjusted relationships in bold.

| | **Mean** | **Lower 95% CI** | **Upper 95% CI** |
|---|---|---|---|
| **(Intercept)** | 29.1698 | 19.1228 | 44.2284 |
| **log(size2017)** | 1.0120 | 0.9680 | 1.0569 |
| **% GBM** | **1.0016** | **1.0001** | **1.0031** |
| **median age, 2023** | **0.9906** | **0.9822** | **0.9991** |
| **% Van. Coastal** | 1.0006 | 0.9980 | 1.0033 |
